# Two Distinct Dynamic Process Models of COVID-19 Spread with Divergent Vaccination Outcomes

**DOI:** 10.1101/2021.04.09.21255166

**Authors:** Ernie Chang, Kenneth A. Moselle

## Abstract

Kinematic models of contagion-based viral transmission describe patterns of events over time (e.g., new infections), relying typically on systems of differential equations to reproduce those patterns. By contrast, agent-based models of viral transmission seek to relate those events or patterns of events to causes, expressed in terms of factors (parameters) that determine the dynamics that give rise to those events.

This paper is concerned with the dynamics of contagion-based spread of infection. Dynamics that reflect time homogeneous *vs* inhomogeneous transmission rates are generated *via* an agent-based infectious disease modeling tool (CovidSIMVL - github.com/ecsendmail/MultiverseContagion). These different dynamics are treated as causal factors and are related to differences in vaccine efficacy in an array of simulated vaccination trials. Visualizations of simulated trials and associated metrics illustrate graphically some cogent reasons for not effectively hard-coding assumptions of dynamic temporal homogeneity, which come ‘pre-packaged’ with the mass action incidence assumption that underpins typical equation-based models of infection spread.

## MOTIVATION – MODELING TEMPORALLY HETEROGENEOUS TRANSMISSION DYNAMICS AND VACCINATION OUTCOMES

### Mass action incidence

This paper reports on work that is concerned with the relationship between vaccination outcomes for SARS-CoV-2 and the possibly heterogeneous dynamics that may determine viral spread in the real-world contexts where vaccination strategies are enacted. In the work reported, both vaccination schedules and transmission dynamics are treated as factors that can vary.

Classic equation-based models of contagion-based spread of infection employ systems of differential equations to describe kinematic features of events (e.g., infections, hospitalizations) over time. These models typically operate under the mass-action assumption, where all infectives within a population are equally likely to make contact with susceptibles. Because transmission takes place within a physical space, mass action models also carry the assumption that all members of the population are equally dispersed within the spaces in which transmission takes place. Transmission dynamics (kinetics and associated measures, e.g., reproduction numbers) are determined in this model on the basis of the concentration of the ‘reactants’ (susceptibles and infectives).

The mass action ‘law’ is positioned centrally in computational systems biology. As stated by Voit and associates: *Over the past 150 years, the mass action law has established itself as the most used default model in mathematical modeling and systems biology, to a point where it is considered undisputed truth that needs no further explanation or justification*.^*1*^

The mass action law^2^ can be traced back to work in the area of physical chemistry, where the law aptly characterizes elementary or first-order reactions (no intervening steps). Where there are intervening mechanisms (second-order or higher-order equations) the law can no longer be applied directly, and rate equations can no longer be reliably deduced solely from the properties of the initial set of reactants in many cases they must be determined experimentally.^3,4^

The mass action assumption typically runs counter to fact, at least until an epidemic is well-established and growing in a population at an exponential rate.^5,6^ There are several ways in which the law may not apply.

- First, and importantly, the ‘reactions’ themselves (viral transmission) may not be first-order events, except perhaps in the sense that the “last-mile” in contagion-based transmission is an infective agent in suitably close proximity to a susceptible agent. Thus, for example, if transmission is conditional upon proximity, but proximity is conditional upon an array of intervening variables, e.g., movement within locations or movement across spaces, then a transmission event is not a first-order expression purely of the number of agents and the viral load of infectives.
- Second, protections such as masking have a direct moderating effect on otherwise primary determinants of infection, e.g., viral load in infective agents.
- Further, if movement itself is conditional upon other factors, e.g., role-prescribed behaviour of agents at any particular point in time, then there exist factors that determine movement which then determine whether first-order mechanisms of transmission take place.

Where these second or higher-order mechanisms can be ignored for *pragmatic* reasons, e.g., because rates of community-based transmission are sufficiently high that contemplated policies will be applied to the population as a whole, then equation-based models based on mass action assumptions may be warranted.

However, if the objective is to generate forecasts in the early stages of an outbreak, when mass action assumptions cannot be justified, or if localized protections are contemplated that are keyed precisely to those second order or higher order factors, then models that embody stochasticity and spatial/temporal heterogeneity are typically recommended or prescribed as alternatives.^7^ A large literature on temporal heterogeneity in transmission dynamics supports this view. See for example the work of Ross and associates.^8^

As well, if the objective is to produce models that reflect most closely the real-world dynamics that govern spread, to support individuals in their efforts to gauge their risk in situations and make informed decisions about the potential ramifications of their actions in the context of their own localized pandemic situation, then models must be developed that reflect those local dynamics.

### Mass action incidence models and virtual clinical trials

Mass action incidence models in effect treat the dynamics underling viral spread as a constant. As such, they are not well-suited to studies that are concerned with vaccination outcomes related to *variation* in transmission dynamics.

This paper is focused on outcomes that could be differentially impacted by temporal heterogeneity in the dynamics governing viral transmission. Our objective is to show how variation in the dynamics that govern viral transmission impact on vaccine effectiveness. Mass action is treated as one target dynamic for simulation but is not regarded as universally or invariably true.

The agent-based modeling tool used for these studies – CovidSIMVL^9^ – is designed specifically to enable simulations where transmission dynamics can vary over time *via* parameters that effectively treat mass action spread as an emergent characteristic of those parameters, without precluding the possibility that other parameters could engender different, possibly temporally heterogeneous dynamics marked by slope and curvature discontinuity. Our motivation is to demonstrate why a mass action assumption should not be regarded as a universal feature of contagion-based viral transmission dynamics. Methodologically, our motivation is to demonstrate how key parameters in an agent-based modeling tool, CovidSIMVL can be configured to generate simulated epidemics that embody different transmission dynamics.

For details on how CovidSIMVL “thinks about” viral transmission (what it assumes, what it does not assume, where key constructs such as “reproduction number” fit into the CovidSIMVL “world-view”) see ***CovidSIMVL Supplementary Material, Sections 1-4***.

## MANIFESTATIONS OF TRANSMISSION DYNAMICS IN COVIDSIMVL

Heterogeneity and stochasticity in viral transmission may be reflected in various features of an infectious disease epidemic, including:

- The topology of transmission chains (e.g., long narrow chains with few secondary infections *vs* broad and shallow trees with a rapidly proliferating number infectious agents transmitting in parallel);^10^
- Average time between infections at different stages in an epidemic;
- Topographical features of viral spread – patterns of diffusion of infective agents through a field populated by a spatially-distributed array of susceptibles.

## “PARTICLE” *VS* “WAVE” DYNAMICS

To examine the effect of variation in transmission dynamics on vaccine effectiveness, we employ an agent-based simulation modeling tool - CovidSIMVL. Through configuration of key parameters as described in the Methods section, we generate two intentionally quite distinct dynamics which, for ease of expression, we term “PARTICLE” and “WAVE”:

- The PARTICLE dynamic enables spatially discontinuous movement among agents. It may begin with a single infective agent in a population of susceptibles, but rapidly evolves into well-mixed array of infectives *via* discontinuous jumps. This evenly dispersed phase in the simulated epidemic corresponds to the idealized model of mass action incidence model spread.
- The WAVE dynamic is reflected in dynamics of spread that remain localized and spatially continuous, resulting in radial spread from an initial location that is surrounded by susceptibles, or lateral spread when initiated at an edge, across a ‘field’ of susceptibles.

## COVIDSIMVL

The CovidSIMVL agent-based simulation system is a flexible generator of model Covid-19 epidemics. CovidSIMVL characterizes contagion-based viral transmission as a process determined by physical proximity of susceptible and infective agents. Transmission rates are determined by three factors:

1. Viral temporal dynamics (e.g., duration of periods of infectivity), keyed to viral load. Zi He and associates,^11^ based on their studies in Wuhan, provide the timings for these periods.
2. Proximity of agents within a finite space (termed a CovidSIMVL “Universe”).
3. Movement of agents within a Universe.

The Multiverse version of CovidSIMVL (not used for the trials reported in this paper) also enables agents to move between different Universes (e.g., school, home, work, restaurant). Agents may display different degrees of movement within different Universes in a Multiverse simulation.

Movement and changes to viral load are subject to stochastic variation

Note that reproduction rate is not a parameter that determines transmission in CovidSIMVL. It is an emergent characteristics of transmission dynamics determined by the three sets of factors set out directly above.

In order to generate simulations that illustrate two distinct transmission dynamics (PARTICLE and WAVE) two parameters are used (detailed in the METHODS section below). The first (“MingleFactor” or “mF) determines what proportion of the total field (a CovidSIMVL Universe) is traversed by any given agent in any given move. The second determines the likelihood that a move will bring agents into sufficiently close proximity for transmission to occur (the “HazardRadius” or “HzR” parameter).

To create a PARTICLE dynamic, HzR is set to a low value, while mF is set to a high value. As a result, agents can move well past a nearest neighbor from one Generation to the next. However, because HzR is set to a low value, moves must result in close proximity for transmission to occur. This prevent high rates of agent movement from generating an explosive growth in transmission, before vaccine effects possibly associated with temporally dynamic heterogeneity are manifested.

By contrast, for the WAVE dynamic, infective agents only need to be in the general vicinity of susceptibles for transmission to occur (HzR is set to a high value). However, agents do not move very far at each iteration. Thus, within a local area containing infective agents, there is a high probability that susceptible agents will become infective, but that zone of infectivity only moves slowly across the space occupied by the full array of susceptibles. This is a dynamic that is more suitable for simulations concerned with localized infections within a space (Universe) or in simulations concerned with localized cross-over transmission from select potentially high-risk spaces (e.g., bar or pub Universes) to other spatially-localized populations, e.g., persons in long-term care Universes.

For details on how CovidSIMVL can be configured to depict transmission in a single Universe, or in a functionally interconnected array of Universes (a CovidSIMVL “Multiverse”) see ***CovidSIMVL Supplementary Material, Section 6***.

## SUB-STUDY #1 – WAVE *VS* PARTICLE DYNAMICS

## METHODS

### The CovidSIMVL Context

#### Agent States

CovidSIMVL is an agent-based model that generates simulated epidemics over the course of iterations (“Generations” in the CovidSIMVL methodology). See ***Figure 1*** for an illustration of snapshot view of an iteration in CovidSIMVL simulated outbreak. As in all CovidSIMVL visualizations, agents are colour-coded according to the following scheme:

**Figure 1.**
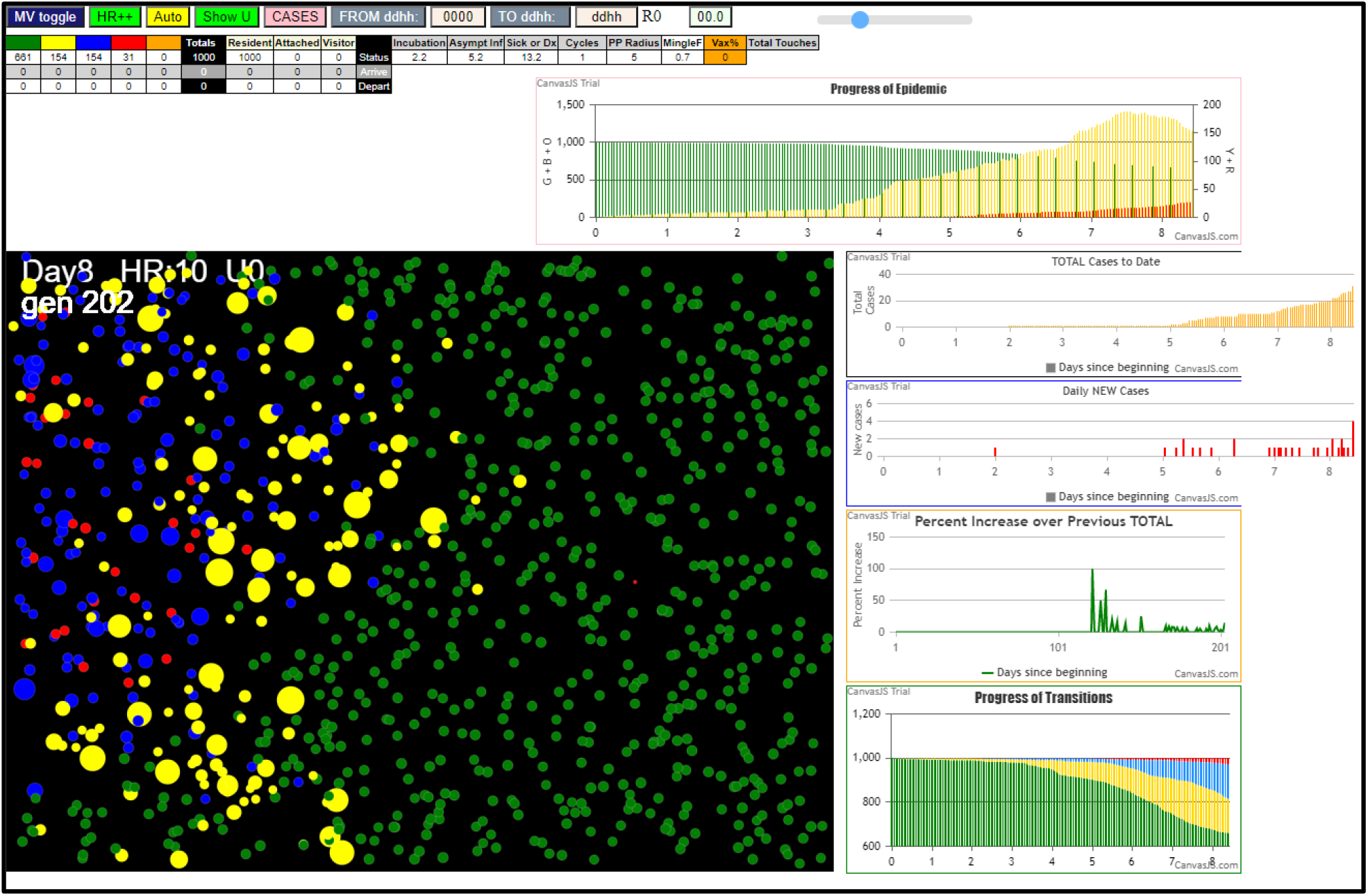
CovidSIMVL Simulated Contagion-Based Epidemic – Single Universe

- Green = Susceptible;
- Yellow = Incubating;
- Blue = Infective but asymptomatic;
- Red = Infective and symptomatic;
- Orange = Inert (recovered and no longer susceptible or deceased or quarantined).

#### CovidSIMVL Spatio-Temporal Frame of Reference

##### Space

each Universe is populated at any given iteration by a collection of agents. Because the Universes are delimited spaces, the number of agents within a Universe translates into density. Because each agent also has a ‘size’ (that varies stochastically), density of agents translates into proximity, which in turn translates into potential for transmission events, because transmission occurs in CovidSIMVL when proximity thresholds are crossed.

##### Time

CovidSIMVL is an agent-based model that generates simulated epidemics over the course of iterations (“Generations” in the CovidSIMVL terminology). “Generations” functions as a clock and may be mapped onto real-time clocks. One hour real-world **time** for each generation is the default configuration for CovidSIMVL for any scheduled set of moves that is intended to reflect movements of people in real-world settings (e.g., 8 generations for a staff person working an 8-hour shift in a simulation of a workplace epidemic).

##### Duration of Infectivity

this is coded in terms of Generations. For example, the “RedDays” parameter in CovidSIMVL represents the number of days that an agent is both infectious and symptomatic.

##### Movement

at each iteration, agents move within a Universe, or they may relocate to a different Universe (e.g., from “Home” to “Workplace”). Movement within a Universe is stochastically varying, according to a Pareto “random walk” distribution. Relocation of agents from one Universe to another is deterministic, according to a pre-determined schedule.

For details on the CovidSIMVL spatio-temporal frame of reference, see ***CovidSIMVL Supplementary Material, Sec. 6*.*1***.

### Configuration of CovidSIMVL to simulate different infectious disease transmission dynamics

Dynamics are configured in CovidSIMVL *via* rules that are organized into a three-fold hierarchy.

1. <u>Primary rules</u> – these are intended to embody physiological determinants of viral spread. They are captured in CovidSIMVL viral temporal dynamics, manifested most directly as the amount of time infected agents spend in different states (incubating; asymptomatic infective; symptomatic infective). Some of these rules are fixed and others can vary. Specifically,
  a. The incubation period and the asymptomatic infective period are set to fixed values, based on a model of viral temporal dynamics reflecting the work of Xi He et al.^12^
  b. The period of Symptomatic Infectivity (“RedDays”) may be set to a particular value at the initiation of a simulation to reflect type/level of protections that would have the effect of removing infective persons from circulation (e.g., testing and quarantine). As well, it can be changed mid-simulation to reflect changes in protections for an epidemic that is mid-course.
2. <u>Secondary Rules</u> – these determine whether the potential for transmission is actualized as the simulation progresses. There are two secondary rules used in the work reported in this paper to generate the WAVE and PARTICLE transmission dynamics:
  a. ***MingleFactor*** (***mF***) – this determines how far an agent moves as the simulation engine cycles through Generations. The amount of movement is subject to stochastic variation. The distance covered in any given move determines the likelihood that an agent will move to a location close to a nearest neighbor *vs* a more distally positioned agent.
  b. ***HazardRadius*** (***HzR***) – this is a delimited area (subject to stochastic variation) surrounding an agent. Transmission can only occur when the areas surrounding a susceptible agent and an infective agent overlap.
  Likelihood that a transmission event will take place as the simulation cycles is a function of the likelihood that a move (whose magnitude is determined by mF) will bring agents into sufficiently close proximity (determined by HzR) for transmission to occur.
3. <u>Tertiary Rules</u> – in CovidSIMVL Multiverse simulations, moves between Universes are set deterministically by Schedules. These schedules also contain potentially Universe-specific adjustments to the mF and HzR values associated with agents. Tertiary rules determine when these moves occur. In other words, they enable different transmission dynamics to be enacted by the same agents when they are re-positioned in different Universes.

#### Events

Equation-based models describe the kinematics of transmission events. Agent-based models such as CovidSIMVL generate the events. Events in CovidSIMVL may be separated into two classes:

1. Events that are not associated with a change of state of an agent, though an agent’s physical location may change within a Universe.
2. Sentinel events – where an agent changes state. These may be divided into two sub-classes:
  a. State changes that are a direct consequence of passage of time in CovidSIMVL time. For example, an agent will transition from an infective state to an inert state after a specified period of time has occurred, regardless of any repositioning of agents over Generations.
  b. State changes that are a direct consequence of movement – if an agent who is susceptible moves into adequately close proximity to an infective agent (Blue or Red) their state changes to Yellow (Incubating).

### WAVE vs PARTICLE Dynamics

**WAVE** – this is defined conceptually as a dynamic that has the following properties:

1. Agents only cover a small amount of space in any given move. A relatively large number of Generations are required for an infection to spread across a field of Susceptibles.
2. HzR set to a level whereby the epidemic does not self-extinguish before it has had a chance to propagate across the field – and across Generations in order to illustrate possibly heterogeneity in dynamics as the infection spreads.
3. RedDays (infectivity) set to a level that, together with HzR, ensures that the epidemic does not self-extinguish.

**PARTICLE** – this is defined conceptually as one that has the following properties:

1. Agents cover sufficient distance in any move to reposition them well beyond the hazard radii set by nearest neighbors.
2. HzR is set to a level whereby the epidemic does not self-extinguish before agents in different states have achieved some approximation of full dispersion across the field.
3. RedDays is set to a level that, together with HzR, ensures the epidemic does not self-extinguish before full dispersion has been achieved.

## RESULTS

### PARTICLE Dynamics

The parameters for the PARTICLE trial were set to the following:

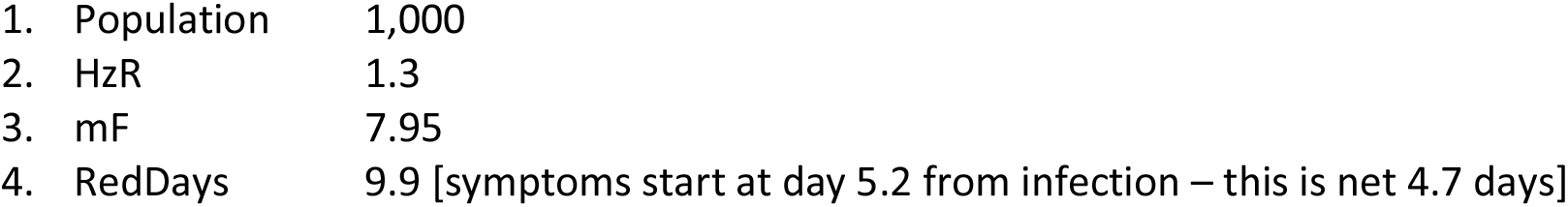

These values were set through trial and error to fully infect (with stochastic margins of error) the entire population. By setting the mF high, and keeping the HzR low, the simulation processes cause agents to move in proportionately large jumps within the virtual arena. Thus, contacts and infections more closely resemble the mass action assumptions of equation based epidemiology models.

See ***Figure 2*** for a depiction of the virtual arena associated with the PARTICLE dynamic epidemic generated with parameters as set out above. Visual examination of the space occupied by the agents suggests a random distribution, though the simulation was initialized with only a single infective agent in a single location in the field depicted. This roughly random distribution of agents is consistent with the presumed distribution of real people in an equation-based epidemiological model that assumes mass action incidence, where every person is equally likely to become infected or pass on the infection, and agents from all compartments are equally dispersed.

**Figure 2.**
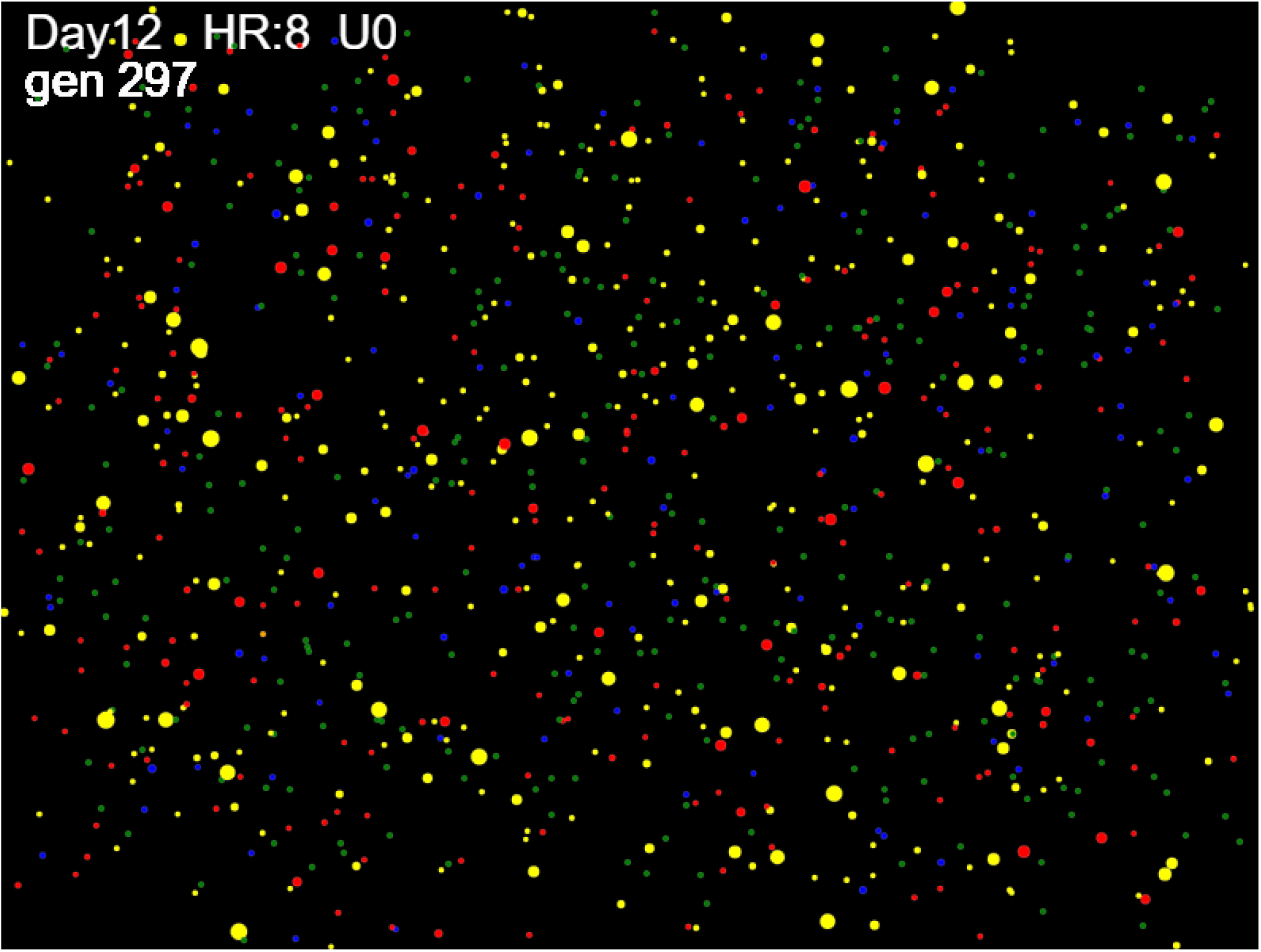
Particle Dynamic – Distribution of Agents

With parameters set at HzR 1.3, RedDays 9.9, and mF 7.95, the trial ended with zero survivors in 1024 Generations. The progression of the epidemic over Generations is depicted in ***Figure 3***. Infectives are depicted in red. The rise is consistent with an exponential growth rate and the fall is consistent with depletion of a supply of Susceptibles.

**Figure 3.**
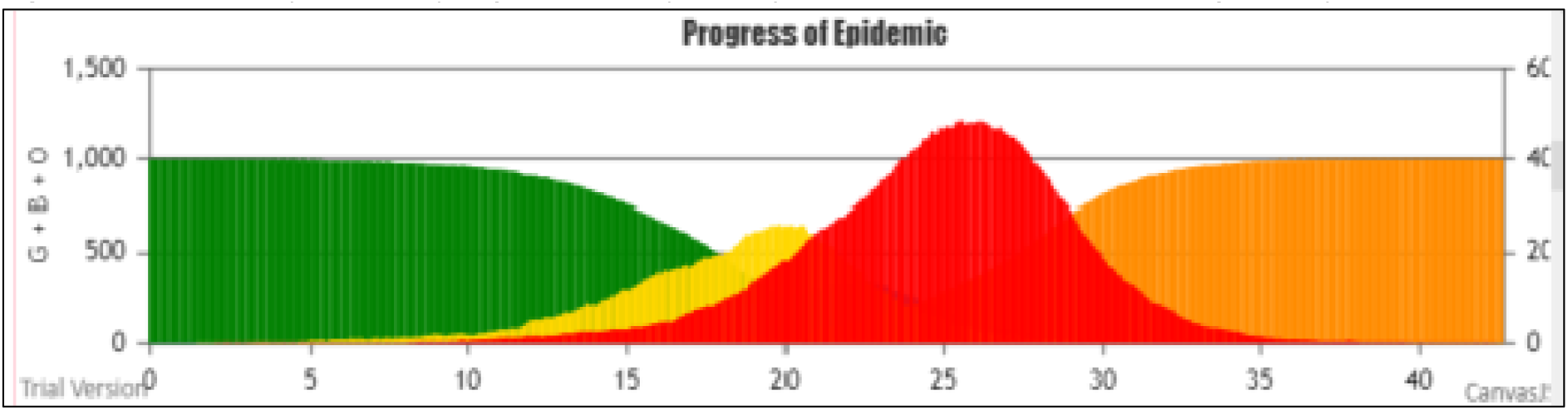
Particle dynamic – progression to point of termination (zero remaining Susceptibles)

See ***Figure 4*** for the growth curve associated with three additional trials produced with the same parameters used to generate ***Figure 3***. The peaks are located at different days but the growth curves are quite similar in shape.

**Figure 4.**
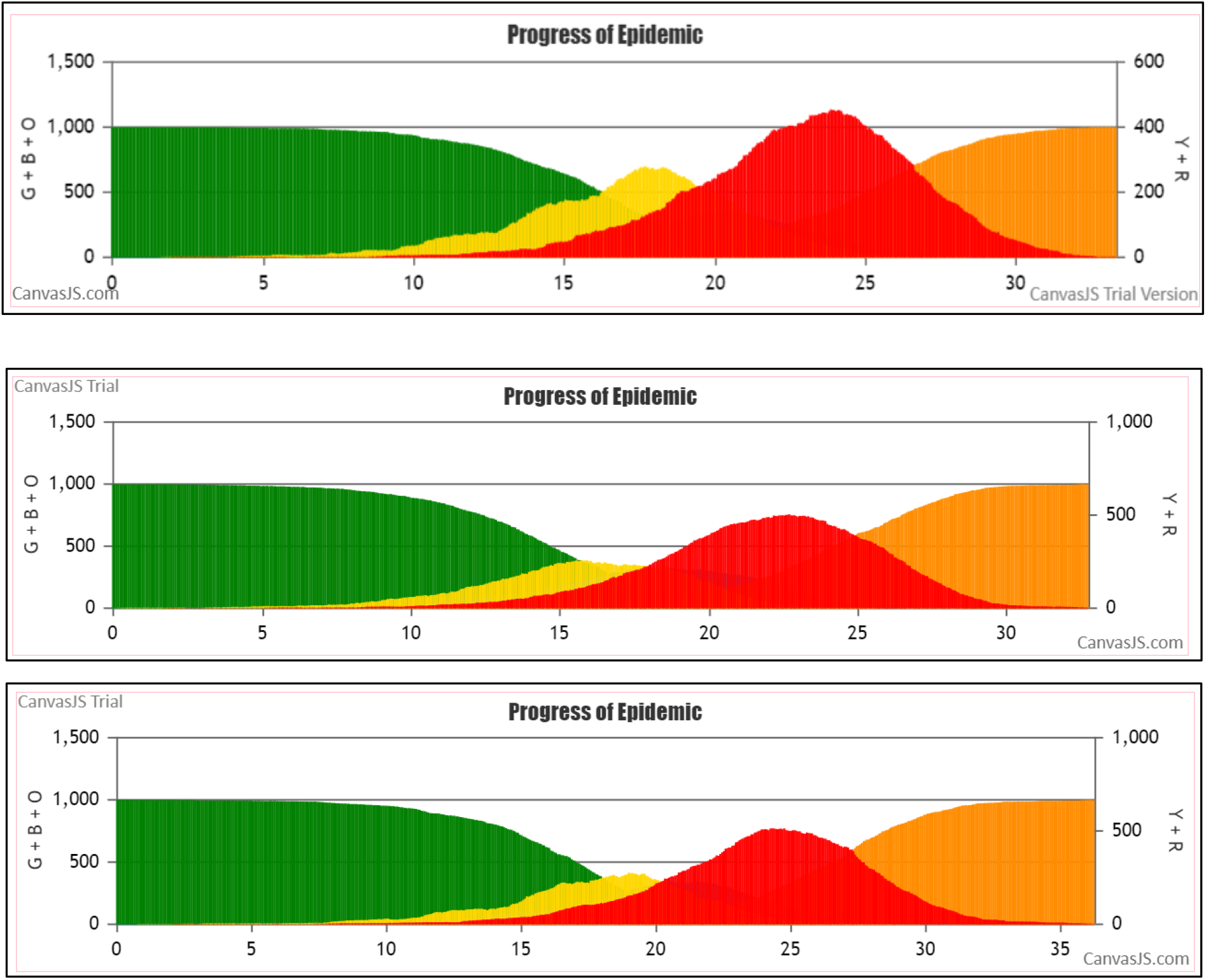
Three additional PARTICLE dynamic simulations.

See ***Appendix II*** for a more detailed longitudinal depiction of a PANDEMIC outbreak in the CovidSIMVL environment.

### Wave Dynamics

For the WAVE trial, the CovidSIMVL parameters were set as follows:

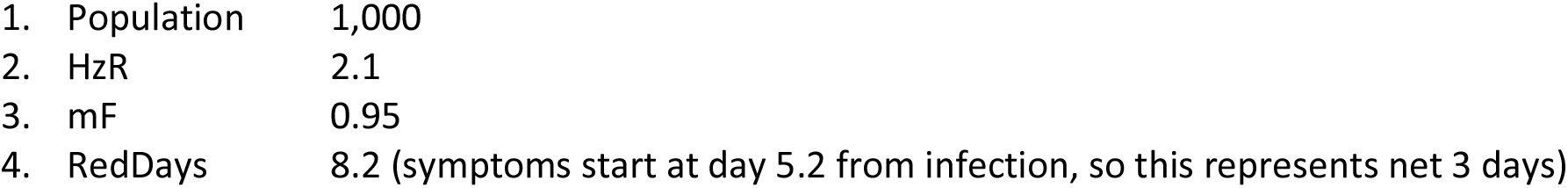

Parameters were set through trial and error to fully infect (with stochastic margins of error) the entire population. The mF value of 0.95 (quite low in the spectrum of mF values used with CovidSIMVL simulations) constrains the agents to remain close to their locality for any given move, despite the stochastic Pareto-like distribution of probable moves. The values on HzR and RedDays ensures that for that degree of movement, the contact rate for infectives does not fall below the level necessary for the epidemic to progress to the point of full infection for the population.

The population of 1,000 Susceptibles (Greens) is distributed randomly at initialization, and we start with a single infected agent, placed randomly. For the WAVE dynamic illustrated in ***Figure 5***, the initial position was in the centre-bottom portion of the Universe. As can be seen in the changes from Generation 243 to 482, the movement of the wave front is coarsely radial. Note that if the position happens to be in a corner, the contagion spreads like a wave-front.

**Figure 5.**
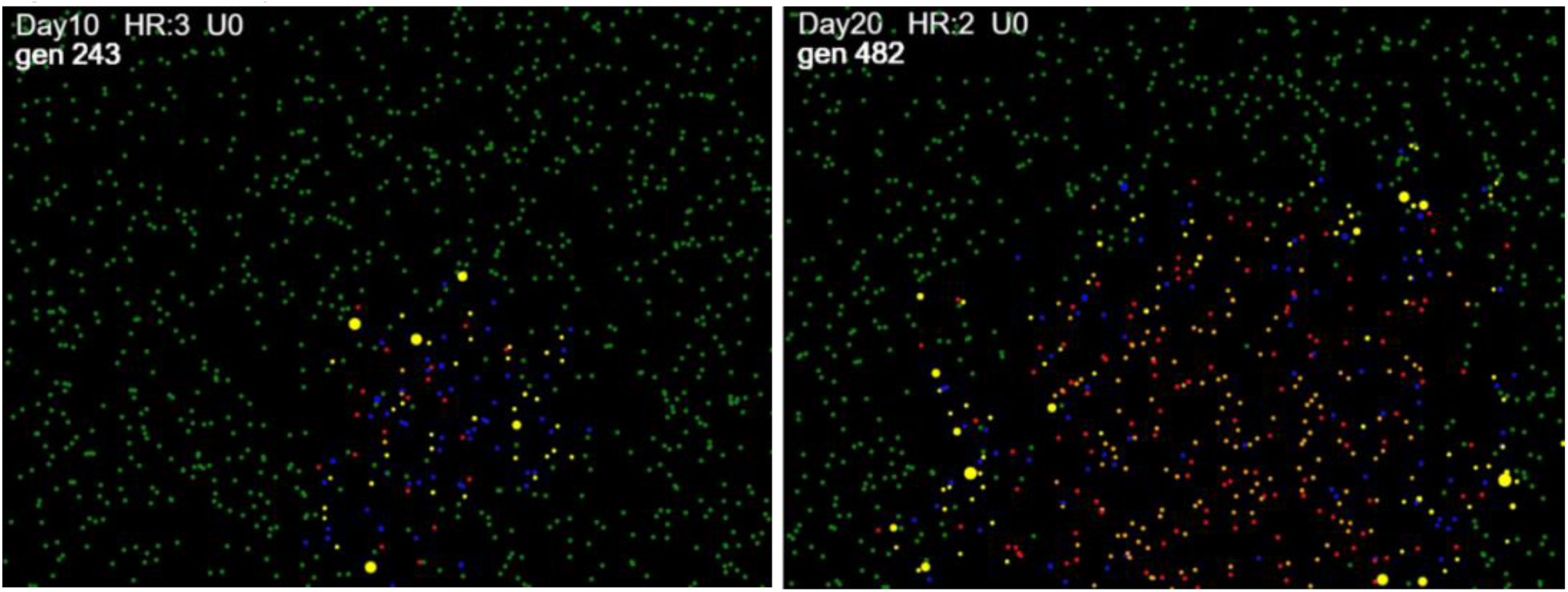
WAVE dynamic

For more detailed depictions of stages in the progress of a Wave dynamic, see ***Appendix 2***

For the epidemic depicted in ***Figure 5***, the trial ended at Generation 1246 (converted to days in the figures, at the rate of 24 Generations per day), at which point there were no remaining infectives and two remaining Susceptibles out of an original population of 1000.

The full progression of the epidemic is depicted in ***Figure 6***. Infectives are depicted in red. In this curve, there appears to be a steady rise in cases until a local maximum is reach on approximately day 13, followed by a plateau of approximately 3 days, followed by a sharp rise and then an extended plateau, followed by the expected drop as the number of available Susceptibles in the finite population is reduced by infection.

**Figure 6.**
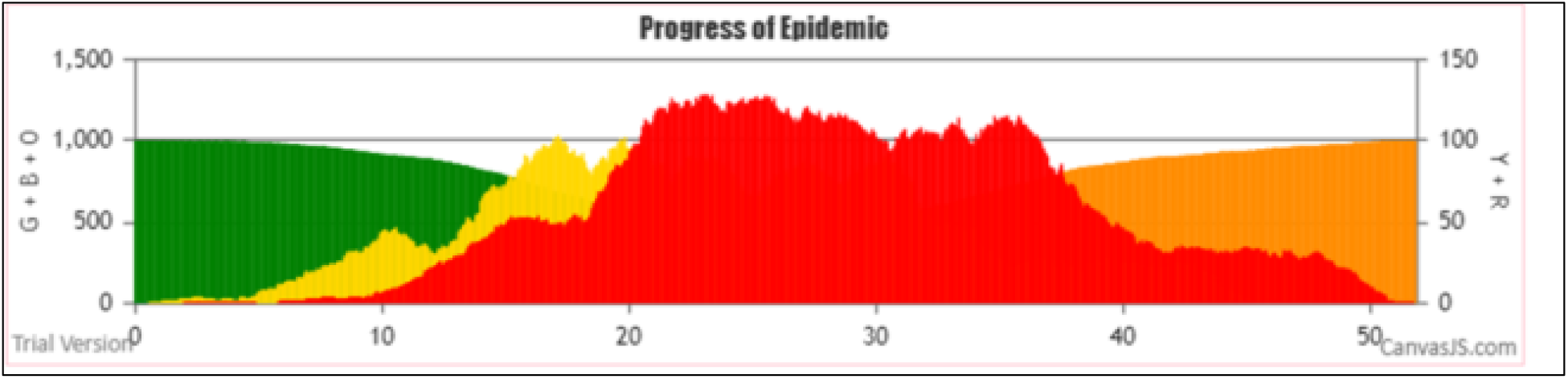
WAVE dynamic – progression to point of termination

Due to stochasticity built into the transmission process *via* variation in the mF and HzR, the same parameters will not generate exactly the same results. Using Wave dynamic parameters, ***Figure 7*** shows a fairly steady rise until roughly day 30, a plateau extending approximately 20 days, and then a sharp reduction due to saturation effects (reduced number of Susceptibles).

**Figure 7.**
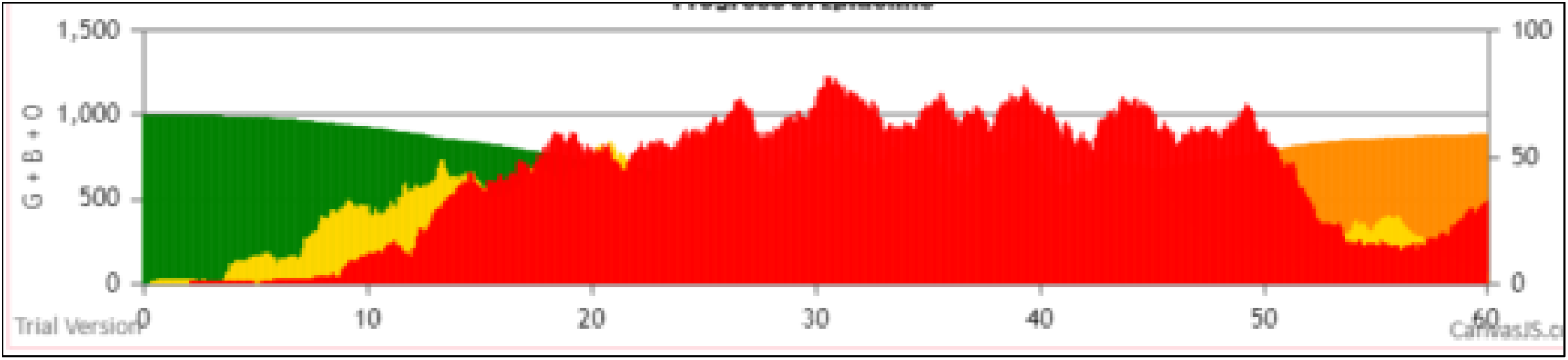
WAVE dynamic – extended plateau

See ***Figure 8*** for three addition Wave simulation, using the same parameters specified above. In the first we see a rise with an initial plateau between days 18 and 24, a gradual rise to a maximum on approximately day 34 and then a drop off in rates as the number of remaining Susceptibles declines. In the second of the two images in ***Figure 8***, we see a rise with a local plateau roughly between days 16 and 18, then a steady rise to a higher rate than the first image in the figure, then a sharp drop-off, followed by an extended plateau between days 28 and 42. That trial terminates at Generation 1253.

**Figure 8.**
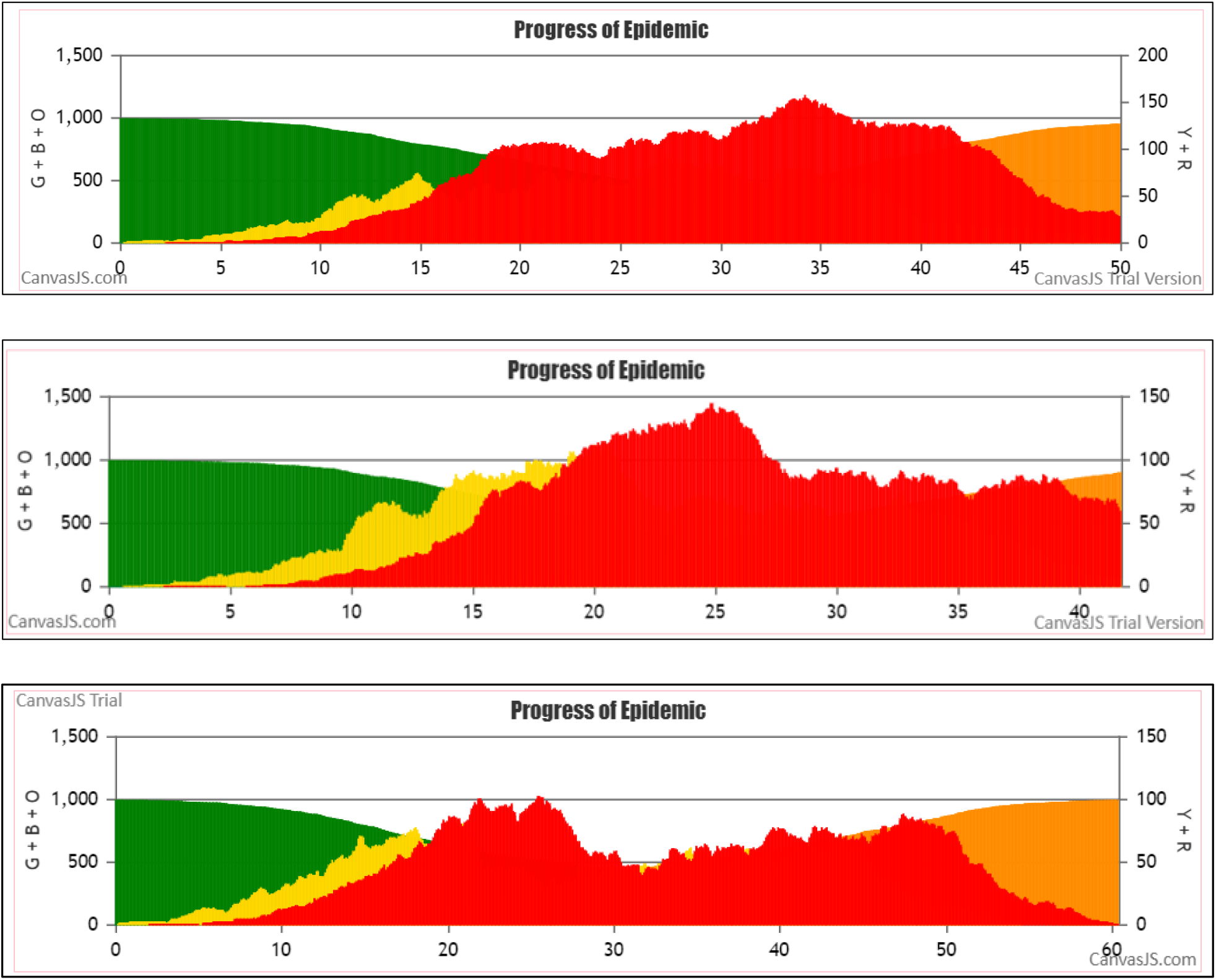
WAVE dynamic – additional trials

In the third trial depicted in ***Figure 8***, there is a fairly steady rise over the course of approximately 20 days, then a fairly sharp drop-off in new infections over the course of about 5 days, then a steady rise over the course of a 20 day period, with a steady decline as all but two remaining Susceptibles are infected. See ***Appendix 2*** for a more detailed longitudinal depiction of this trail.

These curves do not resemble the results obtained by trials set to parameters that generate the PARTICLE dynamic. As well, there is some perhaps noteworthy variation in the shape of these curves over the course of Generations – the dynamic is temporally heterogeneous, marked by multiple slope and curvature discontinuities within trials, and variability between trials. None of the infection points in the curves depicted in ***Figures 6-8*** are caused by mid-course changes to parameters, which are held constant.

### Quantitative Analysis

#### Measuring transmission dynamics

In addition to the usual metrics used to describe the events associated with transmission dynamics (e.g., number of infections at set intervals), CovidSIMVL enables the generation of metrics that quantify features of the dynamics. Dynamic features to be measured include:

1. The proportion of the total population of susceptible agents who become infected over time intervals.
2. The average time between infections at different points in a trial.
3. The topology of transmission chains^13^– for example, is the virus moving through a population *via* a rapidly proliferating, well-dispersed array of infectives or *via* superspreader clusters that are chained together, or *via* long narrow chains with few branching secondary transmissions?

Measures of these dynamic characteristics of a simulated epidemic are derived from contents extracted from the CovidSIMVL *console*.*log* (see ***Figure 9***) which tracks every change of state in an agent. This log includes the identity of the infective agent, the susceptible agent, the location of the infection event (which Universe in a CovidSIMVL multiverse simulation) and which Generation, which provides a time stamp for events. Note references to “***prob = XX***” in *console*.*log*. As explained below, this probability is used to determine whether proximity between a Susceptible and Infective agent gives rise to transmission when an agent has been vaccinated.

**Figure 9.**
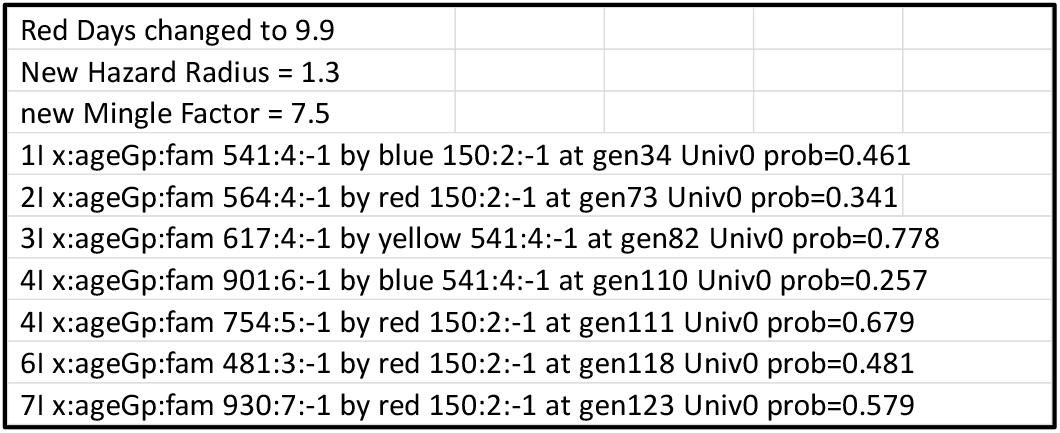
Screen shot – CovidSIMVL console.log.

**Figure 11.**
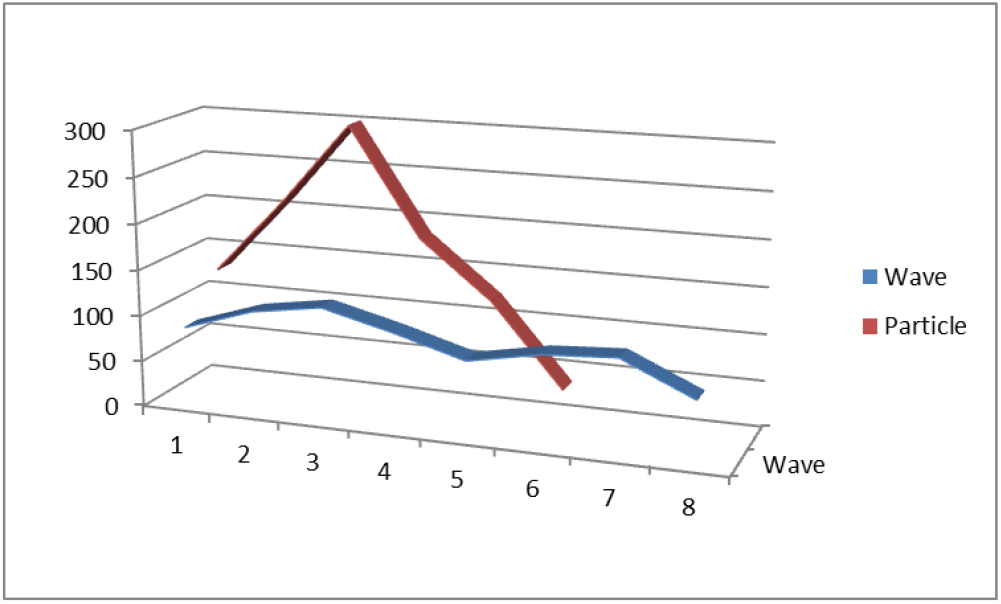
Wave vs Particle dynamics: changes in time-to-next-infection over Generations

#### Metrics derived from CovidSIMVL console.log

The *console*.*log* enables generation of various summary measures:

1. ***theta*** – this is a count of new infections divided by the time period (Generations) during which those new infection events took place. It is a measure of the average elapsed time between new infections. See Moselle & Chang^14^ for details. It corresponds to the construct ““generation time” in classic equation-based modeling. The time period can span the entire simulated epidemic. As well, two different approaches can be used to segmenting the entire outbreak in order to look at possible changes in time between infections over the course of the outbreak:
  a. The epidemic can be segmented on the basis of a specified number of Generations;
  b. The epidemic can be segmented on the basis of a set percentage of the total Susceptible population infected, e.g., first quintile, second quintile.
2. ***R0*** – this is generated directly from a count of secondary infections produced by infected agents up to a specified point in time in a simulated epidemic. In CovidSIMVL, it is an emergent property of the dynamics of transmission. It is not a set value at the time of initialization of a trial, nor is it an estimated quantity based on changes in population-level rates over time.

***Table 1***, below, contains the value of theta at the point when a trial terminates (labelled “theta.all” in the table). The value for WAVE is 1.05 generations between infections, while for PARTICLE it is 0.79 generations between infections – a difference of 38% in favour of longer duration between next infections for the WAVE dynamic. The R0 for WAVE is roughly 5% lower than the R0 for PARTICLE, in keeping with the values for theta.

**Table 1.**
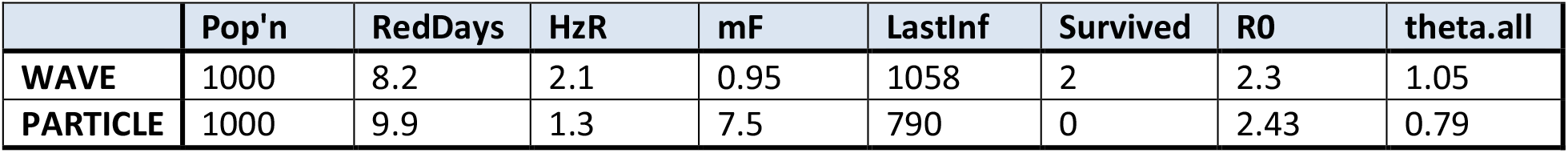
Average time to next infection for Wave and Particle simulations.

|  | Pop'n | RedDays | HzR | mF | LastInf | Survived | R0 | theta.all |
| --- | --- | --- | --- | --- | --- | --- | --- | --- |
| WAVE | 1000 | 8.2 | 2.1 | 0.95 | 1058 | 2 | 2.3 | 1.05 |
| PARTICLE | 1000 | 9.9 | 1.3 | 7.5 | 790 | 0 | 2.43 | 0.79 |

From the chronology of transmission events documented in console.log, we created fixed time frames of 60-Generations, starting at Generation 300 (approximately Day 12.5). Within each frame, we extracted counts of the number of infection events. The results appear in ***Table 2***.

**Table 2.** Dynamic heterogeneity manifesting as changes in time-to-next-infection over Generations.

| Day | 12.5 | 15 | 17.5 | 20 | 22.5 | 25 | 27.5 | 30 |
| --- | --- | --- | --- | --- | --- | --- | --- | --- |
| Gen | From 300 | From 360 | From 420 | From 480 | From 540 | From 600 | From 660 | From 720 |
| WAVE | 85 | 110 | 122 | 103 | 81 | 96 | 101 | 66 |
| PARTICLE | 130 | 211 | 300 | 184 | 123 | 34 |  |  |

This table shows that the PARTICLE processes were more active overall in all but the final 60-Generation frames. The WAVE dynamics gave roughly the same number of infections in each frame, as reflected in the plateau in ***Figure 7***, though a more heterogeneous mix of values would be obtained from the trials in ***Figure 8***.

The relationship between the PARTICLE trial and the WAVE trial in ***Table 2*** are depicted visually in ***Figure*** Because both simulations result in the entire population becoming infected (PARTICLE), or 998 out of 1000 Susceptibles becoming infected (WAVE) almost the same number of infected agents in the peaked PARTICLE curve are distributed over time in the WAVE scenario.

#### Internal Dynamics of WAVE vs PARTICLE

Temporal heterogeneity in transmission dynamics can also be illustrated in CovidSIMVL by segmenting the course of the epidemic on the basis of number of Susceptibles infected. See ***Table 3***. In this table, we show values for ***thetaNNN*** where NNN is a set number of infected agents for a given segment. thetaNNN is computed by dividing time (measured in Generations) by the number of infection events in an interval.

**Table 3.** Dynamic heterogeneity manifesting as changes in time-to-next-infection over fractions of Susceptibles.

|  | theta100 | theta200 | theta300 | theta400 | theta500 | theta600 | theta700 | theta800 | theta900 | theta1000 | Gen1066 |
| --- | --- | --- | --- | --- | --- | --- | --- | --- | --- | --- | --- |
| WAVE | 2.58 | 0.84 | 0.51 | 0.54 | 0.65 | 0.62 | 0.68 | 0.58 | 0.91 | 2.11 | - |
| PARTICLE | 2.68 | 0.43 | 0.32 | 0.28 | 0.23 | 0.17 | 0.20 | 0.22 | 0.35 | 1.18 | 2.20 |

For example, to compute theta100, we take the first 100 infections, identify the time (Generations) of the first and last to determine the total elapsed time for this first 100 infection events. Theta100 is computed by dividing number of Generations required for the first 100 infection events by the number of infection events in an interval.

*Console*.*log* supplies the required data.

Each value of thetaNNN is the time/per infection, and the larger it is, the slower the epidemic. Since we start with only one infected person, the epidemic has to create more simultaneous infectors earlier in the course of the spread, so in both cases theta100 is a relatively large number.

In the case of WAVE, the values for thetaNNN fall consistently between 0.5 and 0.7 for the portion of the simulation falling between theta300 and theta800. This contrasts with PARTICLE. In keeping with the roughly Gaussian distribution of cases over Generations, as depicted in ***Figures 3*** and ***4***, thetaNNN in ***Table 3*** decreases to a low of 0.17 at theta600 and increases on each side, with theta values smaller (faster) than the comparable WAVE thetaNNN values in the same intervals.

These different ways of summarizing the emergence of new infections over the course of the WAVE *vs* PARTICLE dynamics all converge around a conclusion that the PARTICLE dynamic reflects exponential growth with saturation in a roughly randomly dispersed population of finite size. That is, the dispersion of agents rapidly converges on a mass action equilibrium state and the dynamics are in keeping with what must invariably arise with a reproduction number greater than 1 in a population of finite size.

By contrast, the WAVE dynamic is not well described by a Gaussian distribution. Several different runs with the WAVE dynamic parameters highlight time-dependent heterogeneity/discontinuity with local periods of exponential growth punctuated by extended periods of relatively flat reproduction despite an overall reproduction number greater than 1 and no addition of Susceptibles or Infectives from an external reservoir.

**SUB-STUDY #2 – AGE-STRUCTURED POPULATION AND VACCINE OUTCOMES ASSSOCIATED RELATED TO PARTICLE *VS* WAVE DYNAMICS**

## METHODS

CovidSIMVL permits the system to partition the total population of agents into age-groups, and to implement vaccination schedules for a percentage of an age group in a Universe. In this section of the paper, we explore the effects of WAVE vs PARTICLE dynamics on vaccination outcomes in a single fixed Universe.

The population of 1,000 has been tailored to reflect morbidity due to Covid in British Columbia (BC), using 2020 Covid-19 data from the British Columbia Centre for Disease Control, for a period covering January 15, 2020 to December 12, 2020. In ***Table 4***, we calculated the new column “Cases% AgeGp” by taking the “Cases n” for each AgeGroup as a percentage of the “BC Popn” by AgeGroup.

**Table 4.** Distribution of SARS-CoV-2 Cases in BC by Age*

| Age Group | BC Popn | BC % | Cases n | Cases% Total | Case% AgeGp | Hospital | Hosp% Cases | ICU n | ICU% Cases | Death n | Death% Cases |
| --- | --- | --- | --- | --- | --- | --- | --- | --- | --- | --- | --- |
| <10 | 469,351 | 9.1% | 1,736 | 3.99% | 0.37% | 21 | 1.21% | 0 | 0.00% | 0 | 0.00% |
| 10-19 | 527,805 | 10.3% | 3,944 | 9.05% | 0.75% | 20 | 0.51% | 0 | 0.00% | 0 | 0.00% |
| 20-29 | 697,691 | 13.6% | 9,914 | 22.76% | 1.42% | 103 | 1.04% | 13 | 0.13% | 0 | 0.00% |
| 30-39 | 735,052 | 14.3% | 7,986 | 18.33% | 1.09% | 197 | 2.47% | 37 | 0.46% | 1 | 0.01% |
| 40-49 | 646,035 | 12.6% | 6,506 | 14.94% | 1.01% | 215 | 3.30% | 53 | 0.81% | 8 | 0.12% |
| 50-59 | 718,272 | 14.0% | 5,685 | 13.05% | 0.79% | 340 | 5.98% | 102 | 1.79% | 17 | 0.30% |
| 60-69 | 673,131 | 13.1% | 3,539 | 8.12% | 0.53% | 402 | 11.36% | 141 | 3.98% | 46 | 1.30% |
| 70-79 | 435,062 | 8.5% | 2,067 | 4.75% | 0.48% | 491 | 23.75% | 153 | 7.40% | 124 | 6.00% |
| 80-89 | 187,443 | 3.6% | 1,418 | 3.26% | 0.76% | 385 | 27.15% | 59 | 4.16% | 262 | 18.48% |
| 90+ | 49,726 | 1.0% | 763 | 1.75% | 1.53% | 132 | 17.30% | 7 | 0.92% | 220 | 28.83% |
| Total | 5,139,568 | 100.0% | 43,558 | 100.00% | 0.85% | 2,306 | 5.29% | 565 | 1.30% | 678 | 1.56% |

\*Covering January 15, 2020 to December 12, 2020.

We use the column “Case%AgeGp” column to construct a new table of 1,000 cases:

1. We fix the total “Cases n” at 1,000.
2. We use the “Cases % Total” to fill the new “Cases n” rows for each age group. Thus, for age group <10, from “Cases % Total” of 3.99% X 1,000 (total in “Cases n”) we get 40, and so on.
3. We use the derived “Cases% AgeGp” to calculate the BC population. So, if 40 cases make up, from “Case% AgeGp”, 0.37% of the BC population, the BC population for the age group <10 would be 40/0.37% or 10,775, and so on for the other age groups.

The results appear in ***Table 5***, which distributes the 1,000 patients into age-groups using “Cases n”. This table is useful as the age structure is the same as the morbidity structure reported, but for 1,000 cases. Working from these figures, we can compare simulated results for infection events to BC reported obtained values for Cases, Hospitalizations, ICU Admissions, and Deaths.

**Table 5.**
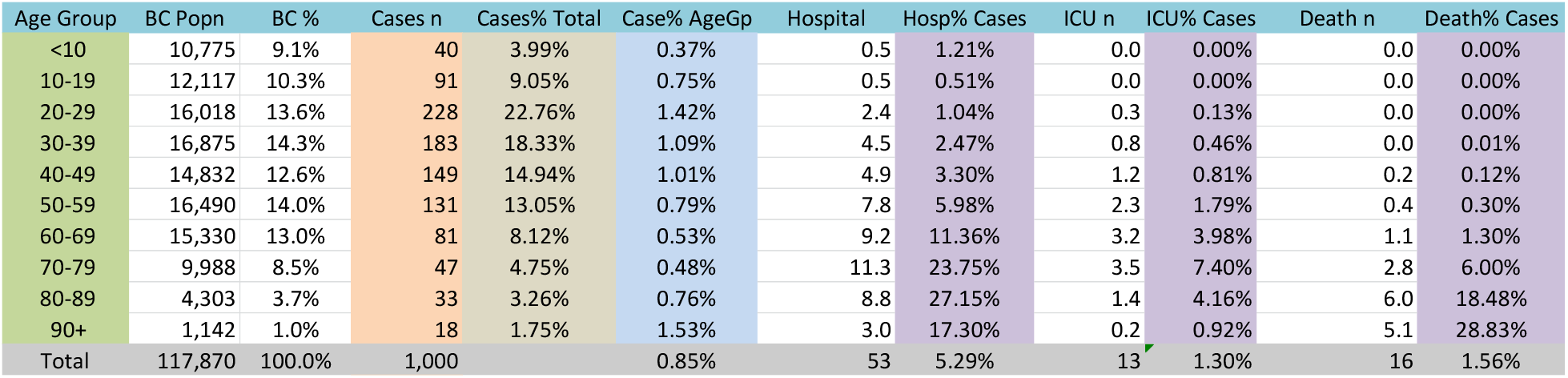
CovidSIMVL population of 1000 agents fitting the age distribution of cased in BC

| Age Group | BC Popn | BC % | Cases n | Cases% Total | Case% AgeGp | Hospital | Hosp% Cases | ICU n | ICU% Cases | Death n | Death% Cases |
| --- | --- | --- | --- | --- | --- | --- | --- | --- | --- | --- | --- |
| <10 | 10,775 | 9.1% | 40 | 3.99% | 0.37% | 0.5 | 1.21% | 0.0 | 0.00% | 0.0 | 0.00% |
| 10-19 | 12,117 | 10.3% | 91 | 9.05% | 0.75% | 0.5 | 0.51% | 0.0 | 0.00% | 0.0 | 0.00% |
| 20-29 | 16,018 | 13.6% | 228 | 22.76% | 1.42% | 2.4 | 1.04% | 0.3 | 0.13% | 0.0 | 0.00% |
| 30-39 | 16,875 | 14.3% | 183 | 18.33% | 1.09% | 4.5 | 2.47% | 0.8 | 0.46% | 0.0 | 0.01% |
| 40-49 | 14,832 | 12.6% | 149 | 14.94% | 1.01% | 4.9 | 3.30% | 1.2 | 0.81% | 0.2 | 0.12% |
| 50-59 | 16,490 | 14.0% | 131 | 13.05% | 0.79% | 7.8 | 5.98% | 2.3 | 1.79% | 0.4 | 0.30% |
| 60-69 | 15,330 | 13.0% | 81 | 8.12% | 0.53% | 9.2 | 11.36% | 3.2 | 3.98% | 1.1 | 1.30% |
| 70-79 | 9,988 | 8.5% | 47 | 4.75% | 0.48% | 11.3 | 23.75% | 3.5 | 7.40% | 2.8 | 6.00% |
| 80-89 | 4,303 | 3.7% | 33 | 3.26% | 0.76% | 8.8 | 27.15% | 1.4 | 4.16% | 6.0 | 18.48% |
| 90+ | 1,142 | 1.0% | 18 | 1.75% | 1.53% | 3.0 | 17.30% | 0.2 | 0.92% | 5.1 | 28.83% |
| Total | 117,870 | 100.0% | 1,000 |  | 0.85% | 53 | 5.29% | 13 | 1.30% | 16 | 1.56% |

### Vaccination Mode in CovidSIMVL

In Chang & Moselle (2020)^15^, we use the term “modes” to refer to two different *de facto* administration schedules (one dose *vs* both doses) for the Pfizer and Moderna vaccines. These modes were defined in relationship to the two dose schedules set out in the EUA (Emergency Use Authorization) submissions of the manufacturers.^16,17^ We also incorporated into the simulated vaccine trials a vaccine we termed “Hybrid” which had a second dose scheduled at 45 days. Effectiveness rates for Moderna, Pfizer and were set to correspond to documentation supplied by the manufacturers (Pfizer, Moderna). “Hybrid” effectiveness was set to fall in a range roughly in the “middle” of various published estimates of Pfizer and Moderna effectiveness.

The vaccines are assumed to be inactive for 14 days with no protection, then with 75% protection till day 28, after which the person has 95% protection (upon administration of the second dose). The vaccine’s efficacy becomes operational in a simulation at the time a contact has been detected between a viral carrier and a Susceptible. If the random number drawn is greater than the threshold of protection, the viral transfer occurs, even if the Susceptible has been vaccinated. If the threshold is not exceeded, there is no state change for the agents.

### Implementing Vaccination Schedules in CovidSIMVL

Every agent is randomly assigned a probability score ranging between 0 and 1, which is used to determine whether they become infected following vaccination, when positioned suitably proximally to an infective agent. The value assigned to any given agent shows up in *console*.*log* with the notation “prob=.*value”*. This probably score is a threshold value used to determine whether a vaccinated Susceptible will become infected when movement puts that agent in proximity to an Infective. If the value of ***prob*100*** exceeds a specified rate for vaccine effectiveness that has been coded into CovidSIMVL, then a Susceptible will become infected. For example, a vaccinated Susceptible with a randomly assigned probability of .8 will become infected if a move positions them proximally to an Infective agent, so long as the vaccine in the simulation has an effectiveness rate set to a value lower then 80%.

The vaccinated person is able to transmit at the same level as the protection given to a Susceptible. In other words, if the random number drawn by the Susceptible is greater than the threshold (say 95%) the vaccinated transmitter may infect.

The console.log produces a trace of encounters involving vaccinated or unvaccinated Susceptibles and infective agents, and it records the result of the encounter, both for vaccinated Infectives and for Susceptibles.

In the version of CovidSIMVL used in this paper, functionality has been expanded to enable vaccination to take place at any point in a simulation, for any specified portion of any given age group. However, for all of the work reported in this paper, all vaccinations are administered to targeted agents at the same time. Administration shows up in the console.log records as a vaccination event, with a listing of identifiers for all vaccinated agents.

### “Standard” vs “Fast Forward” Modeling of Vaccine Outcomes

The vaccine temporal model as described above and implemented in Chang & Moselle^18^ entails no protection for the first 14 days. We call this the **STANDARD** approach. A second set of trials are reported in this paper using what we term the **FAST FORWARD** “FF” approach, in which the date of first injection is recorded as 14 days prior to the ***current date***, so that effectively, the second injection, and associated protection, takes place at the time of initiated of the trial (Generation = 0). Note that in the STANDARD trials that follow, the vaccinations are given at time T=0.

### Configuration of CovidSIMVL to achieve PARTICLE vs WAVE dynamics

Different vaccine outcome trials were carried out in the context of simulations where the parameters were set to create WAVE dynamics or PARTICLE dynamics. The parameters set out in Study #1 were employed. To reiterate:

|  |  |  |  |
| --- | --- | --- | --- |
| WAVE | RedDays=8.2 | HzR=2.1 | mF=0.95 |
| PARTICLE | RedDays=9.9 | HzR=1.3 | mF=7.5 |

Again, recall that the value set for RedDays are the value of days from the time of initial infection, and onset of symptoms is at 5.2 days after initial infection. Consequently, the duration of RedDays=8.2 is 3 days after onset.

### Age Stratification

Age Groups for vaccination trials are set as follows:

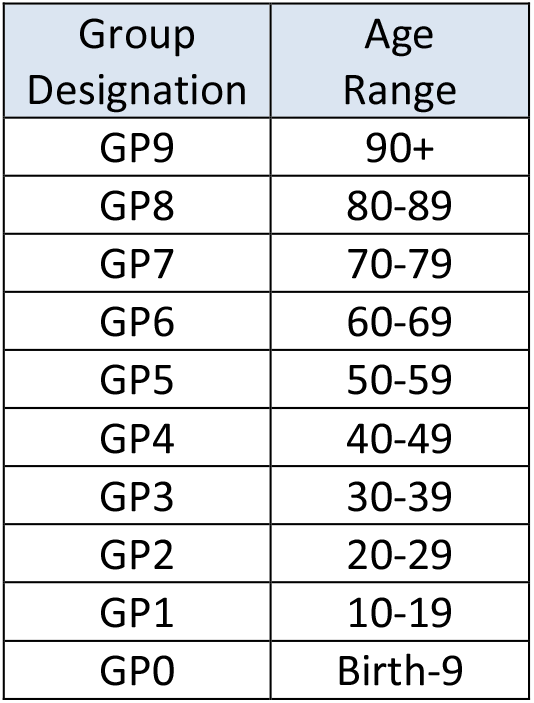

| Group Designation | Age Range |
| --- | --- |
| GP9 | 90+ |
| GP8 | 80-89 |
| GP7 | 70-79 |
| GP6 | 60-69 |
| GP5 | 50-59 |
| GP4 | 40-49 |
| GP3 | 30-39 |
| GP2 | 20-29 |
| GP1 | 10-19 |
| GP0 | Birth-9 |

### Vaccination Schedules

The vaccination schedules are as follows:

Vaccination Schedules The vaccination schedules are as follows:
| Schedule Designation | 100% Vaccinated | 50% Vaccinated |
| --- | --- | --- |
| Schedule 0 | None | None |
| Schedule 1 | 9, 8, 7 (98 'persons') |  |
| Schedule 2 | 9, 8, 7 (98 'persons') | 4, 3, 2 (560 'persons'; 280 vaccinated) |
| Schedule 3 | 5, 4, 3, 2 (691 'persons') |  |

We conceive of these simulated vaccine trials as follows:

<u>Schedule 0</u> – this is a no-vaccination control.

<u>Schedule 1</u> – older adults only - this considers protection of all older adult populations, though subject to the limits of effectiveness of the vaccines, but no vaccination for potential sources of infection for the largely but not fully protected older adults.

<u>Schedule 2</u> – partial protection of fully vaccinated older adult – this considers the effects of fully vaccinating the older adults but only partially vaccinating the populations that might transmit into these older adult groups.

<u>Schedule 3</u> – this tests the influence of suppressing transmitters alone on risks to the elderly by vaccinating agents in the age range 20-59, which represents 70% of the population of 1000 agents.

The trials terminate when no further infections are possible. The final status of all age groups is produced below for WAVE and PARTICLE.

In summary, then, we start with the **Standard** approach (14 days of no protection, then 75% to day 28, when 95% protection is conferred with administration of the second dose.). We compare the results of Schedule 1, 2 and 3 for WAVE and PARTICLE processes.

We then repeat these trials using the **Fast Forward** approach.

## RESULTS

### Standard approach

For these trials, the assumption is no protection for the first 14 days, the 75% from day 14-28, and 95% protection with the second dose.

Using the *console*.*log* as source data for the trials, the results are shown in ***Table 7***, below,

**Table 7.** Standard approach, survival by age group for three vaccination schedules

| STANDARD APPROACH |  |  | Terminate | Survivors | R0 | Gp9 | Gp8 | Gp7 | Gp6 | Gp5 | Gp4 | Gp3 | Gp2 | Gp1 | Gp0 |
| --- | --- | --- | --- | --- | --- | --- | --- | --- | --- | --- | --- | --- | --- | --- | --- |
| WAVE | Sched 0 | Trial 1 | 1246 | 2 | 2 | 0/18 | 0/33 | 0/47 | 1/81 | 0/131 | 0/149 | 0/182 | 2/228 | 0/91 | 0/40 |
|  | Sched 1 | AVG | 1443 | 34 | 2 | 6/18 | 11/33 | 15/47 | 2/81 | 1/131 | 0/149 | 0/182 | 1/228 | 0/91 | 0/40 |
|  | Sched 2 | Trial 1 | 1706 | 182 | 2 | 4/18 | 18/33 | 27/47 | 1/81 | 1/131 | 31/149 | 39/182 | 60/228 | 2/91 | 0/40 |
|  | Sched 3 | Trial 1 | 2381 | 481 | 2 | 1/18 | 7/33 | 7/47 | 11/81 | 90/131 | 88/149 | 116/182 | 143/228 | 13/91 | 6/40 |
| PARTICLE | Sched 0 | Trial 1 | 1024 | 0 | 2 |  |  |  |  |  |  |  |  |  |  |
|  | Sched 1 | AVG | 902 | 14 | 2 | 3/18 | 4/33 | 7/47 | 0/81 | 0/131 | 0/149 | 0/182 | 1/228 | 0/91 | 0/40 |
|  | Sched 2 | Trial 1 | 904 | 93 | 2 | 6/18 | 12/33 | 11/47 | 0/81 | 0/131 | 15/149 | 18/182 | 0 | 0/91 | 0/40 |
|  | Sched 3 | Trial 1 | 1105 | 390 | 2 | 0/18 | 1/33 | 1/47 | 0/81 | 79/131 | 81/149 | 92/182 | 134/228 | 3/91 | 0/40 |

For the WAVE dynamic, the key observations are:

1. For Schedule 0 (no vaccination control) all but two Susceptibles become infected by the time the trial self-terminates (no more infectives). This is by design – the simulated was calibrated (RedDays, HzR, mF) to ensure that it ran to full or near-full infection of the population.
2. Schedule 3 - full vaccination of persons ranging from 20-59 (GP 2, 3, 4, 5) produced the expected effect:
  a. For the vaccinated groups – partial but not complete protection, due to the 14 days of transmission before the vaccine took effect, to the partial suppression from days 14-28, and to substantial but not 100% complete suppression following the second dose.
  b. For the unvaccinated groups (60 and older) – very limited protection for the elderly (shown in orange in ***Table 7***).
3. Schedule 1 (only older adults vaccinated) *vs* Schedule 2 (older adults vaccinated, 50% adults aged 20-49 vaccinated) – older adults are better protected overall when a significant fraction of the adults are also vaccinated. Again, this is the expected result.

For the PARTICLE trials, the pattern is similar, though the epidemic progresses more quickly, and as such, the number of protected individuals is smaller. For example, in Schedule 2, the total number of survivors (uninfected agents) when the trial terminated was 93 for the PARTICLE dynamic and 182 for the WAVE dynamic. For the older adults aged 70 and above, there were 49 uninfected agents out of the total population of 98. By contrast, for the PARTICLE dynamic, there were 29 out of 98 uninfected agents for the same vaccine schedule.

***Tables 8*** and ***9*** show differences in the speed at which the epidemic progresses in the WAVE *vs* PARTICLE vaccination trials. Recall that theta is the time between subsequent transmissions, expressed in terms of Generations, within a time frame. Here, theta.all is the average time between transmission based on total elapsed time from the first to last infection. It reflects the average separation between any given transmission within a trial. In ***Tables 8*** and ***9***, theta for WAVE is consistently higher than for PARTICLE.

**Table 8.** Standard Approach - time-between-infections (rate of infection) for WAVE dynamics

| WAVE | Trial | theat.al |
| --- | --- | --- |
| Schedule 0 | Trial 1 | 1.05 |
|  | Trial 1 | 1.11 |
|  | Trial 2 | 1.20 |
|  | Trial 3 | 1.19 |
|  | Trial 4 | 1.57 |
|  | Trial 5 | 1.27 |
| Schedule 2 | Trial 1 | 1.79 |
| Schedule 3 | Trial 1 | 4.20 |

**Table 9.** Standard Approach - time-between-infections (rate of infection) for PARTICLE dynamics

| PARTICLE | Trial | theat.al |
| --- | --- | --- |
| Schedule 0 | Trial 1 | 0.76 |
| Schedule 1 | Trial 1 | 0.68 |
|  | Trial 2 | 0.66 |
| Schedule 2 | Trial 1 | 0.65 |
| Schedule 3 | Trial 1 | 1.32 |

### Standard approach - Survival – WAVE vs PARTICLE

For the Standard approach, ***Table 10*** summarizes the results relating specifically to vaccine effectiveness, appearing in various locations above, highlighting differences between PARTICLE and WAVE dynamics.

**Table 10.** Survival under different vaccination schedules within WAVE vs PARTICLE DYNAMICS

| Vaccination Schedule | # 'Persons' Vaccinated | PARTICLE Dynamic |  | WAVE Dynamic |  |
| --- | --- | --- | --- | --- | --- |
|  |  | # Survivors | Survivors/ Vaccinated | # Survivors | Survivors/ Vaccinated |
| Schedule 0 | 0 | 0 |  | 2 |  |
| Schedule 1 | 98 | 14 | 14.3% | 34 | 34.7% |
| Schedule 2 | 378 | 93 | 24.6% | 182 | 48.1% |
| Schedule 3 | 691 | 390 | 56.4% | 481 | 69.6% |

The overall survival rate for the WAVE dynamic is higher than the rate for the PARTICLE dynamic, regardless of the vaccination schedule. However, even in the best case scenario simulation (Schedule 3, WAVE dynamic) approximately 1/3 of the agents become infected.

### Fast Forward approach

In the Standard approach, outcomes reflect protection conferred by the vaccine, as well as spread that takes place before the vaccine confers protection, and once that protection starts to take effect.

The “Fast Forward” simulation design for CovidSIMVL evaluates vaccination outcomes for populations where the vaccination is effectively back-dated to 14 days before the start of the trial. This is in keeping with standard epidemiological designs that seek to provide ‘pure’ measures of vaccine effectiveness following a latency period. An example would be a recent major study of vaccine outcomes in the UK,^19^ or the Canadian National Advisory Council on Immunization (NACI) – “*Excluding the first 14 days before vaccines are expected to offer protection, both vaccines showed an efficacy of 92% up until the second dose (most second doses were administered at 19-42 days in the trials)*.^20^

In the Fast Forward CovidSIMVL trials, the vaccine is set to a 75% protection threshold which becomes operational on Generation 1 of the trial (rather than Generation 336, i.e., 14 ‘days’ in the Standard trials, above). The Fast Forward simulations use the same schedules as in the Standard trials. Results appear in ***Table 11***.

**Table 11.** Fast Forward approach, survival by age group for three vaccination schedules

| WAVE |  | Terminate | Survivors | R0 | Gp9 | Gp8 | Gp7 | Gp6 | Gp5 | Gp4 | Gp3 | Gp2 | Gp1 | Gp0 |
| --- | --- | --- | --- | --- | --- | --- | --- | --- | --- | --- | --- | --- | --- | --- |
| Sched 0 | Trial 1 | 1488 | 3 | 2.3 | 0/18 | 0/33 | 0/47 | 0/81 | 0/131 | 2/149 | 0/182 | 2/228 | 0/91 | 0/40 |
| Sched 1 | Trial 1 | 1112 | 54 | 2.31 | 11/18 | 13/33 | 26/47 | 0/81 | 0/131 | 1/149 | 3/182 | 1/228 | 0/91 | 0/40 |
| Sched 2 | Trial 1 | 1591 | 268 | 1.94 | 14/18 | 22/33 | 32/47 | 2/81 | 0/131 | 56/149 | 65/182 | 76/228 | 2/91 | 0/40 |
| Sched 3 | Trial 1 | 2778 | 579 | 1.77 | 0/18 | 2/33 | 4/47 | 8/81 | 114/131 | 112/149 | 156/182 | 176/228 | 6/91 | 4/40 |
| PARTICLE |  |  |  |  |  |  |  |  |  |  |  |  |  |  |
| Sched 0 | Trial 1 | 867 | 0 | 2.43 |  |  |  |  |  |  |  |  |  |  |
| Sched 1 | Trial 1 | 995 | 68 | 2.29 | 14/18 | 23/33 | 29/47 | 0/81 | 0/131 | 0/149 | 1/182 | 2/228 | 0/91 | 0/40 |
| Sched 2 | Trial 1 | 1255 | 294 | 2.1 | 18/18 | 31/33 | 43/47 | 34/81 | 63/131 | 96/149 | 119/182 | 154/228 | 39/91 | 18/40 |
| Sched 3 | Trial 1 | 1962 | 646 | 1.67 | 1/18 | 2/33 | 3/47 | 7/81 | 129/131 | 135/149 | 164/182 | 209/228 | 2/91 | 2/40 |

For the Fast Forward approach, ***Table 12*** summarizes the results relating specifically to vaccine effectiveness, appearing in various locations above, highlighting differences between PARTICLE and WAVE dynamics.

**Table 12.** Survival under different vaccination schedules within WAVE vs PARTICLE DYNAMICS

| Vaccination Schedule | # 'Persons' Vaccinated | PARTICLE Dynamic |  | WAVE Dynamic |  |
| --- | --- | --- | --- | --- | --- |
|  |  | # Survivors | Survivors/Vaccinated | # Survivors | Survivors/Vaccinated |
| Schedule 0 | 0 | 0 |  | 3 |  |
| Schedule 1 | 98 | 68 | 69.4% | 54 | 55.4% |
| Schedule 2 | 378 | 294 | 77.8% | 268 | 70.9% |
| Schedule 3 | 691 | 646 | 93.5% | 579 | 83.8 % |

The key observations are:

1. The times to termination are higher than in the Standard Approach, as would invariably be the case for a vaccine that was indeed conferring any protection.
2. As well, the overall protection is higher than in the Standard Approach, which directly reflects 14 days of unprotected spread in the Standard approach:
  a. For the WAVE dynamic, Schedules 1 *vs* 2 *vs* 3, the survivors are 54 *vs* 268 *vs* 579 for Fast Forward, compared to 34 *vs* 182 *vs* 481 for Standard.
  b. For PARTICLE, the survivors are 68 *vs* 294 *vs* 647 for Fast Forward, compared to 14 *vs* 91 *vs* 390 for Standard.
3. For the Fast Forward trials, the WAVE survivorship is now marginally lower than PARTICLE compared to the Standard approach, where survivorship was higher. For example, for Schedule 1 and Schedule 2, the WAVE *vs* PARTICLE survivors are:
  a. Schedule 1 – WAVE *vs* PARTICLE survivors are 54 vs 68 in the Fast Forward trials. By contrast, in the Standard approach there are more survivors in the WAVE dynamic trials - 34 *vs* 14.
  b. Schedule 2 – there is a similar inversion of the relative survival rates for WAVE *vs* PARTICLE: 268 (WAVE) to 294 (PARTICLE) for Fast Forward, whereas the results were 182 (WAVE) to 93 (PARTICLE).
4. As was shown in the Standard approach, for the Fast Forward approach, the Schedule 3 approach of vaccinating transmitters in Gps 2,3,4,5 does not protect the elderly.

### Standard *vs* Fast Forward approaches – vaccine schedule effectiveness for the Older Adults

In this section, we focus specifically on vaccine outcomes for the total of 98 persons falling in GPS 7,8 and 9 (i.e., 70+ years old). To highlight the impact of the first 14 days of unprotected spread in the Standard trials, we compute the difference between the number of survivors (uninfected) in the older adult group in the Standard vs Fast Forward trials. These figures appear in the columns in ***Table 13*** labeled ““WAVE 14d” and “PARTICLE 14d”. Positive values indicate that there were more survivors in the Fast Forward trials than the Standard trials.

**Table 13.** Vaccine schedule effectiveness for older adults (70+ years old)

| SCHEDULES | WAVE Std | WAVE ff | PARTICLE Std | PARTICLE ff | WAVE 14d | PARTICLE 14d |
| --- | --- | --- | --- | --- | --- | --- |
| Schedule 1 | 32 | 50 | 14 | 66 | 18 | 52 |
| Schedule 2 | 49 | 68 | 29 | 92 | 19 | 63 |
| Schedule 3 | 15 | 6 | 2 | 6 | -9 | 4 |

ff = Fast Forward trials; Std = Standard Trials

For Schedules 1 and 2, both of which involve 100% vaccination of the older adult group, we obtain the expected result – more survivors when we remove the effect of 14 days of no protection in the Standard trials.

For Schedule 3 we obtain a different pattern of results. Recall that for Schedule 3, the older adults are not vaccinated, but younger groups aged 20 to 59 are vaccinated.

1. For the PARTICLE Dynamic, vaccinating the younger groups confers very little protection. There are only 4 out of 98 older adult agents in an uninfected state when the trial self-terminates.
2. However, for the WAVE dynamic, the data show that there are more uninfected older adult agents in the Standard trial than in the Fast Forward trial. In other words, for a Schedule that entails vaccination of younger but not older adults, when we arithmetically remove the effect of 14 days of no protection for all 1000 agents and look at the number of uninfected cases at termination of the trial (no more infectives) we find more uninfected older adults in the Standard trial (with 14 days of no protection) than in the Fast Forward trial (which starts when protections take effect). The effect is pronounced (in comparison to results from the other Schedules) for the WAVE dynamic, and the effect is also moving in the same direction for the PARTICLE dynamic.

We discuss this apparently anomalous results below.

## DISCUSSION

### Dynamics of WAVE vs PARTICLE as determined by degree of agent movement

The overarching motivation of this study is to highlight the effects of transmission dynamics on vaccination outcomes. Because findings arising from a simulation model are not subject to any unmeasured external sources of variance, in order to generate some possible explanations for the apparently anomalous findings noted directly above, we need to focus internally on the kinetic features of WAVE vs PARTICLE spread.

CovidSIMVL entails a Monte Carlo approach to determining physical movement of agents at each Generation in a simulation. The MingleFactor (mF) parameter determines the relative magnitude of movement. Recall that WAVE has parameters of 0.95 for MingleFactor, compared to 7.5 for PARTICLE. This means that the agents in WAVE remain roughly (but not exactly) in the region of a nearest neighbor for any given cycle. By comparison, for the PARTICLE dynamics the agents are likely to move to a region that is proportionately significantly larger than the separation of agents at any given point in time. In other words, the movements are relatively “large”.

For the WAVE dynamic, the relatively small degree of movement results in spread of infection as a virtual wave front, in which infected agents at small distances from the transmitter become the new margins of the epidemic. In comparison, PARTICLE agents are all moving at significant distances and this separation takes them to new neighbors.

This means that transmitting WAVE agents must find new uninfected agents in their neighborhood to keep the epidemic going, and this dynamic is subject to local saturation effects. In contrast, with the PARTICLE agents, each Generation puts infective agents into proximity with a new array of possibly susceptible neighbors. In this dynamic, saturation effects would occur at a field level, not a local level.

This produces the observations that theta.all for Standard WAVE trials are consistently higher than for PARTICLE processes.

### Dynamics affected by vaccinations; vaccination outcomes impacted by transmission dynamics

In CovidSIMVL, vaccination effects are introduced into simulations by randomly assigning a probability score to each agent, ranging between 0 and 1, and evaluating that score against a coded value for vaccine effectiveness. If the probability for a susceptible agent is greater than the value for vaccine effectiveness, transmission occurs, even if the susceptible agent has been vaccinated.

What this means is that the *potential* for transmission emerges as a function of agent movement and resulting proximity or lack thereof. If movement brings an infective agent into proximity with a susceptible agent, the transmission will be “accepted” vs “rejected” depending on the vaccine protection level and a randomly assigned value to the agent. The proportion of accepted *vs* rejected transmissions is related directly to the vaccine effectiveness standard evaluated against the randomly assigned probability assigned to each agent.

As the vaccinated proportion of the population increases, the more likely it is that a move will result in an instance of rejected transmission. As the *console*.*log* shows (in ***Appendix 1***), toward the end of a trial there are many more failures than successes in transmission. In Standard, Susceptible agents are infected quickly in the first 14 days, but in Fast Forward, resistance is immediate. For this reason, the Fast Forward trials require more Generations than Standard trials to reach a point of termination.

In both the WAVE and PARTICLE processes, the presence of many vaccinated also slows the creation of more simultaneous transmitters. Whether in Standard or in Fast Forward approaches, this prolongs the overall time, resulting in an increase in the number of survivors at termination.

The effect of removing the 14-day unprotected infection in the Fast Forward approach accentuates this reduction of simultaneous transmitters, and we would therefore expect that the survivor count goes up, while the total time to termination also increases. This is borne out in the tables above for STANDARD and FAST FORWARD, and in the Charts appearing in ***Figure 12***.

**Figure 12.**
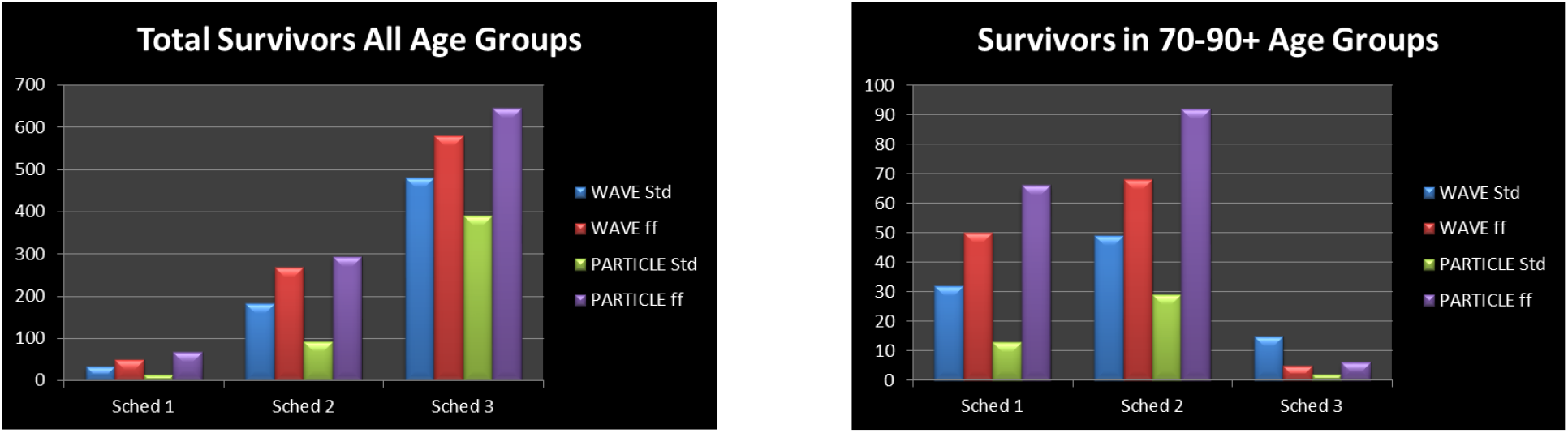
Survivors, WAVE vs PARTICLE under Different Vaccination Schedules, Standard vs Fast Forward.

Schedule 1 vaccinates the Age Groups 90+, 80-89, and 79-79. We see in the 70-90+ chart that indeed WAVE in Fast Forward has more survivors, and PARTICLE in Fast Forward also has more survivors than PARTICLE in Standard approach.

A consistent and perhaps counterintuitive observation is that WAVE in the Standard modeling approach has higher survivorship than PARTICLE in Standard, but WAVE in Fast Forward has consistently *fewer* survivors than PARTICLE in Fast Forward-see blue and green versus red and purple bars in ***Figure 12***, above.

The difference between Standard and Fast Forward is 14 days of unprotected transmission for those who have been vaccinated (Schedule 1 has 98 vaccinated, Schedule 2 has 378, and Schedule 3 has 690). This head start permits the PARTICLE agents to create freely a significant number of transmitters by day 14. On the other hand, transmission within the WAVE dynamic is less advantaged by the opportunity to create simultaneous agents freely in the first 14 days, as the slower, localized neighborhood crawl of the wave front reduces the opportunity to spawn multiple simultaneously infecting agents. We can see this in ***Table 2*** earlier in this paper – at day 12.5 and 15, the Standard approach for WAVE has 85 and 110 infections in the 60 generation time frames, while the PARTICLE dynamic has 130 and 211 transmissions.

However, all of the trials begin with a single Infective agent, regardless of encoded dynamic (PARTICLE *vs* WAVE) or timing of introduction of vaccine effects (STANDARD *vs* FAST FORWARD). Consequently, in the Fast Forward trials, infective agents encounter vaccine-protected agents immediately. In the Standard trials, for Particle, rapid dispersion of agents throughout the field effectively “kick-starts” the epidemic, optimizing the opportunity to create multiple simultaneously and diffusely located infecting agents in the first 14 days. That advantage is lost in the Fast Forward trials, and the loss (relative to the rate of spread in unprotected 14 days) is proportionately greater for the PARTICLE dynamic.

As noted above, the 90+ Chart shows almost no survivors, thus rejecting that the hypothesis that vaccinating transmitters alone is significantly protective for the other age groups.

### Intensity vs Time

In Sub-Study #1, we observed that the WAVE dynamic process is both slower and of longer duration than PARTICLE. The characteristic charts depicting infections (and other changes of state) over time follows a typical Gaussian curve. By contrast, the WAVE charts have a longer base with one or more truncated cones, mixed in with an array of differently-contoured plateaus. The total number of infections is the same for a starting population of 1000, so what this observation implies is that we can trade intensity for time.

### Growth curves

We note that repeat PARTICLE trials produces curves with very similar shapes, differing largely in when the peak number of new cases appears over the course of the simulated epidemic. These epidemics are well described by the typical logistic growth curves that arise with exponential spread of a virus in a population of finite resources (Susceptibles).

For the WAVE dynamic, it is not at all apparent that the curves can be well described with a logistic growth curve. There are periods of exponential growth in some simulations, followed by pronounced flattening or downturns, followed by increases, despite the fact that there is no infusion of new Susceptibles into the simulation, nor is there in infusion of Infectives from an external reservoir, nor are there any mid-stream changes to the parameters that govern the dynamics. Large numbers of these WAVE trials need to be run in order to determine what are modal characteristics of the apparently temporally heterogeneous/discontinuous dynamics that arise in the simulations.

## SUMMARY

In this paper, we have defined two dynamics for simulated contagion processes based on viral temporal dynamics for Covid-19, on the extremes of agent movements that are either relatively local or marked by large displacements. These were named WAVE and PARTICLE. Even when a simulation is initialized with a single infective, large displacements of all agents over the course of iterations within a PARTICLE dynamic rapidly converges on a randomized, evenly spaced dispersion of agents in all states (Susceptible, Incubating, Infective, Inert). This corresponds to the situation embodied in models that conform to a mass action incidence model.

We defined a patient population of 1,000 with age distributions similar to that of the case prevalence of Covid-19 in British Columbia for the period January to December 2020, and generated simulations that demonstrated these two different dynamics. For the PARTICLE trials, with parameterized ‘bio’-dynamics configured to biases odds in favour of a homogeneous mixing of agents, and reproduction at a rate greater than one secondary infection per agent (CovidSIMVL does not actually pre-specify a reproduction number directly), with saturation effects, the simulation produces events over time that trace to an expected logistic curve. For WAVE dynamics, an array of different growth curves were obtained which do not conform to a logistic model. These curves displayed temporal discontinuity/heterogeneity in the dynamics they depicted.

To understand better the kinetics that were producing these differently shaped growth curves (different between PARTICLE and WAVE; different within WAVE from one simulation to another) we abstracted characteristics from the transmission events associated with these two dynamics. The rate of progression of the epidemics was characterized in terms of ***theta***, the time in generations between subsequent infections in a given time frame. We noted the trade-off of time-between-infections and intensity in WAVE.

Because a mass action incidence scenario does not characterize the early stages of an outbreak, and it never characterizes the real-world situations in which transmission occurs (typically a person-to-person event within a bounded real-world context) we took up the question of how different vaccination strategies would impact on survival under different transmission dynamics. To this end, we applied different vaccination schedules to the populations, focusing on protecting the elderly with 100% vaccination, then adding 50% to age groups 20-49. Finally, we tested the hypothesis that vaccinating the most infectious age groups might protect the elderly. That was specified operationally for the simulations as the group ranging in age from 20-59, or 70% of the total simulated population. We modified the vaccination trials to a mode in which we Fast Forward on the time lag for vaccination protection, so that we negated the 14-day wait time after the first injection.

The results of the vaccination trials are discussed in detail above. The summary finding is that that while vaccination brings significant increases in survivors, it is only in Schedule 2 in Fast Forward for WAVE and PARTICLE where we see survivorship > 50% for the elderly age groups.

This low survival rate for the elderly is no doubt due to the rather aggressive dynamics that we set, the reason being that we tried to find the lowest parameters that would ensure an almost complete infection for the 1,000 population, in order to study the effects of vaccination on protection in an active epidemic. We could have configured the trials for lower infection base rates, but the vaccination protection trials would have to remove the effect of “natural survivors” from the lower intensity epidemic, possibly obscuring the effects of vaccination schedules, or transmission dynamics, or the interaction of vaccination schedules and transmission dynamics.

These simulations are scenarios, and their relationship to In Real Life (IRL) is by no means predictive or assured. Nevertheless, this kind of simulation, which technically is a Monte Carlo Markov Chain (MCMC) microsimulation of a clinical trial, permits the elaboration of possibilities and measurements in ways that cannot be realized IRL.

The trials in this study look at infection as the sentinel event, and they evaluate different transmission dynamics and vaccination strategies against that event. The results can easily be carried over to the percentages that are hospitalized, go to ICU or suffer death from the tables generated. Since there are no cost estimates for these downstream events, we leave their elaboration to others.

## Data Availability

The study presents results from an agent-based simulation modeling tool. Aggregated data re Distribution of SARS-CoV-2 Cased in British Columbia by Age are publicly available from the BC Centre for Disease Control (http://covid-19.bccdc.ca/)

https://github.com/ecsendmail/MultiverseContagion

## ACKNOWLEDGMENTS

This work was supported by grant funds from the Michael Smith Foundation for Health Research (COVID-19 Research Response Funding) with co-funding from the Victoria Hospitals Foundation. As well, we acknowledge the Vancouver Island Health Authority (VIHA or “Island Health”) for support from several departments and programs, including the Applied Clinical Research Unit, Research & Capacity Building, and various units within the Innovation, Analytics & Information division. The authors declare no financial conflicts of interest with this work.

## I. APPENDIX

**SHOWING INFECTION PREVENTIONS/FAILURES FOR VACCINATION TRIAL**

At the start of a trial in Fast Forward PARTICLE dynamic with Groups 2,3,4,5 vaccinated at 100%

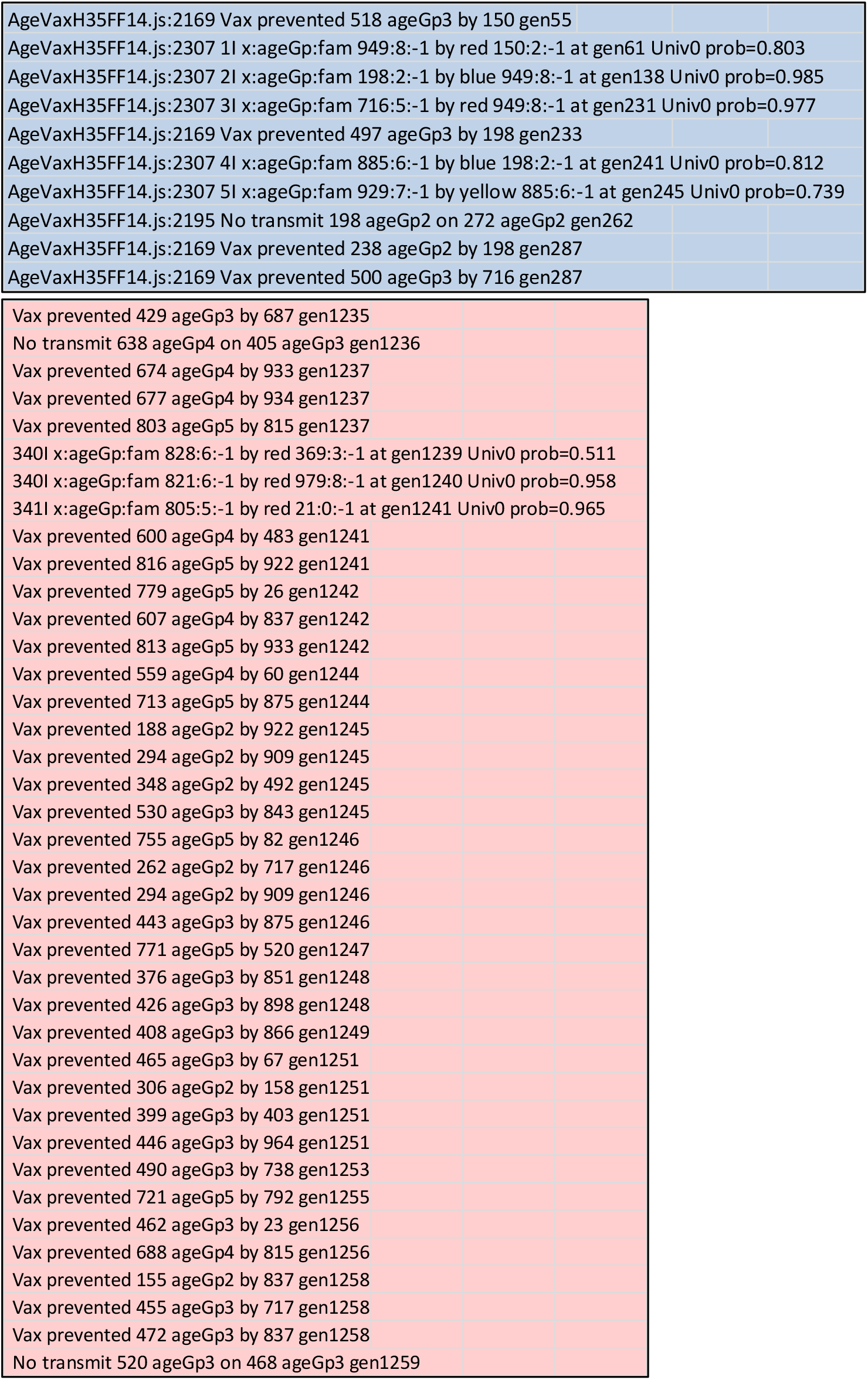

## II. APPENDIX

**PARTICLE, WAVE DYNAMICS – LONGITUDINAL VISUALIZATIONS**

**PARTICLE DYNAMIC** - HzR = 1.3; mF = 7.5; RedDays = 9.9: **Generations 1 – 450**

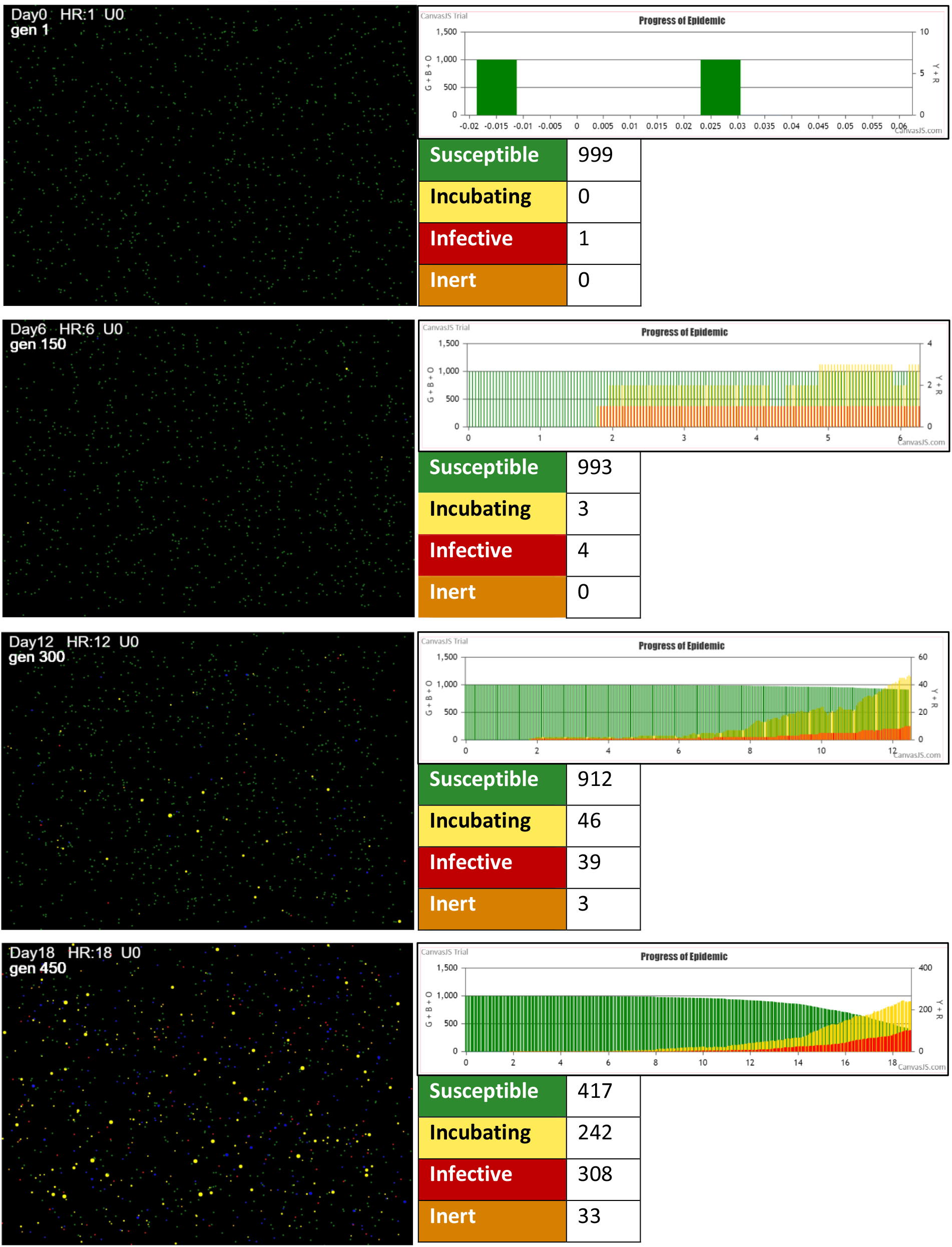

**PARTICLE DYNAMIC** - HzR = 1.3; mF = 7.5; RedDays = 9.9: **Generations 600 – TERMINATION**

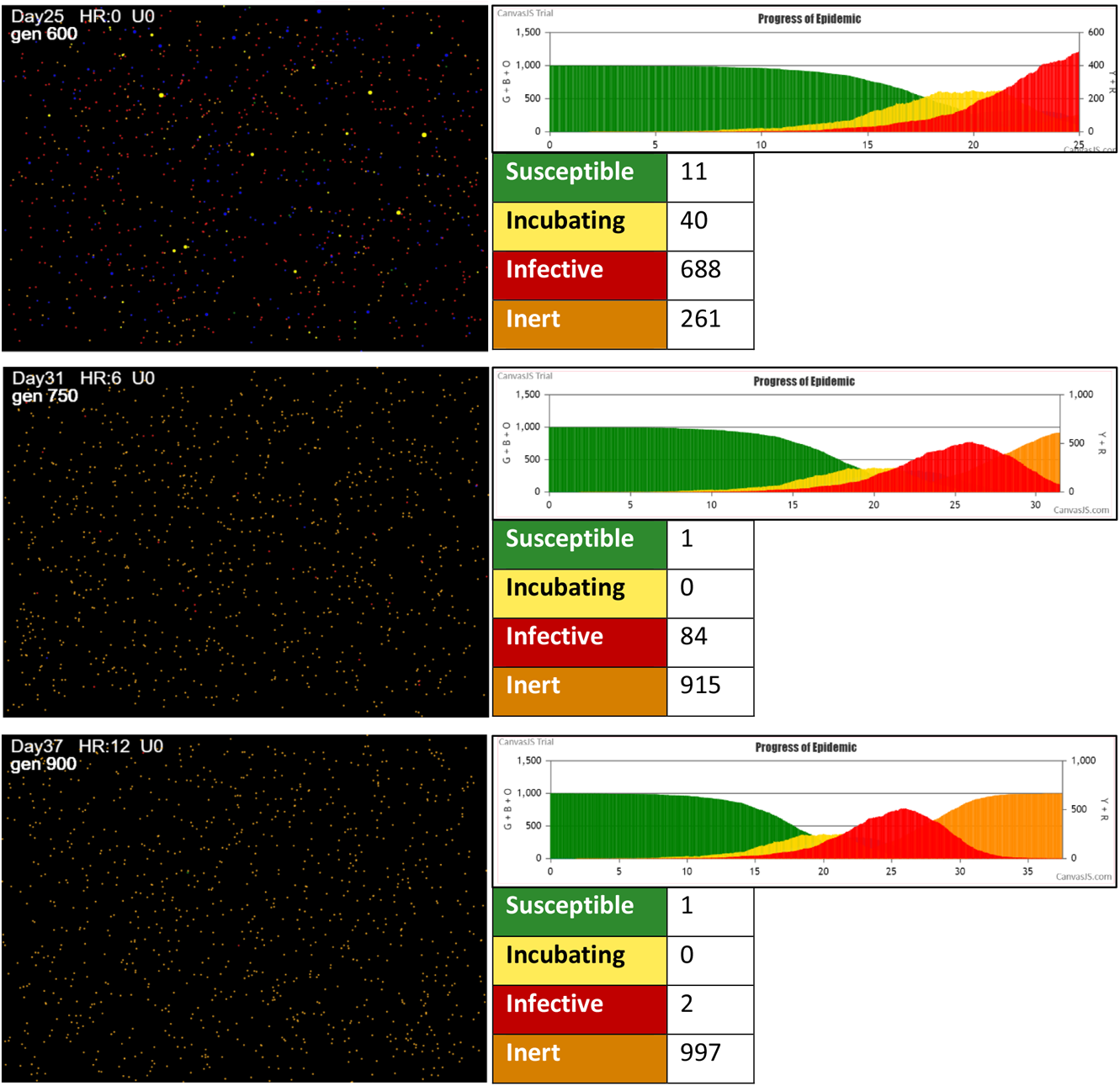

**Terminates at Generation 909 with zero remaining Susceptibles**.

**WAVE DYNAMIC** - Parameters HzR = 2.1; mF = 0.95; RedDays = 8.2 **GENERATIONS 1 – 450**

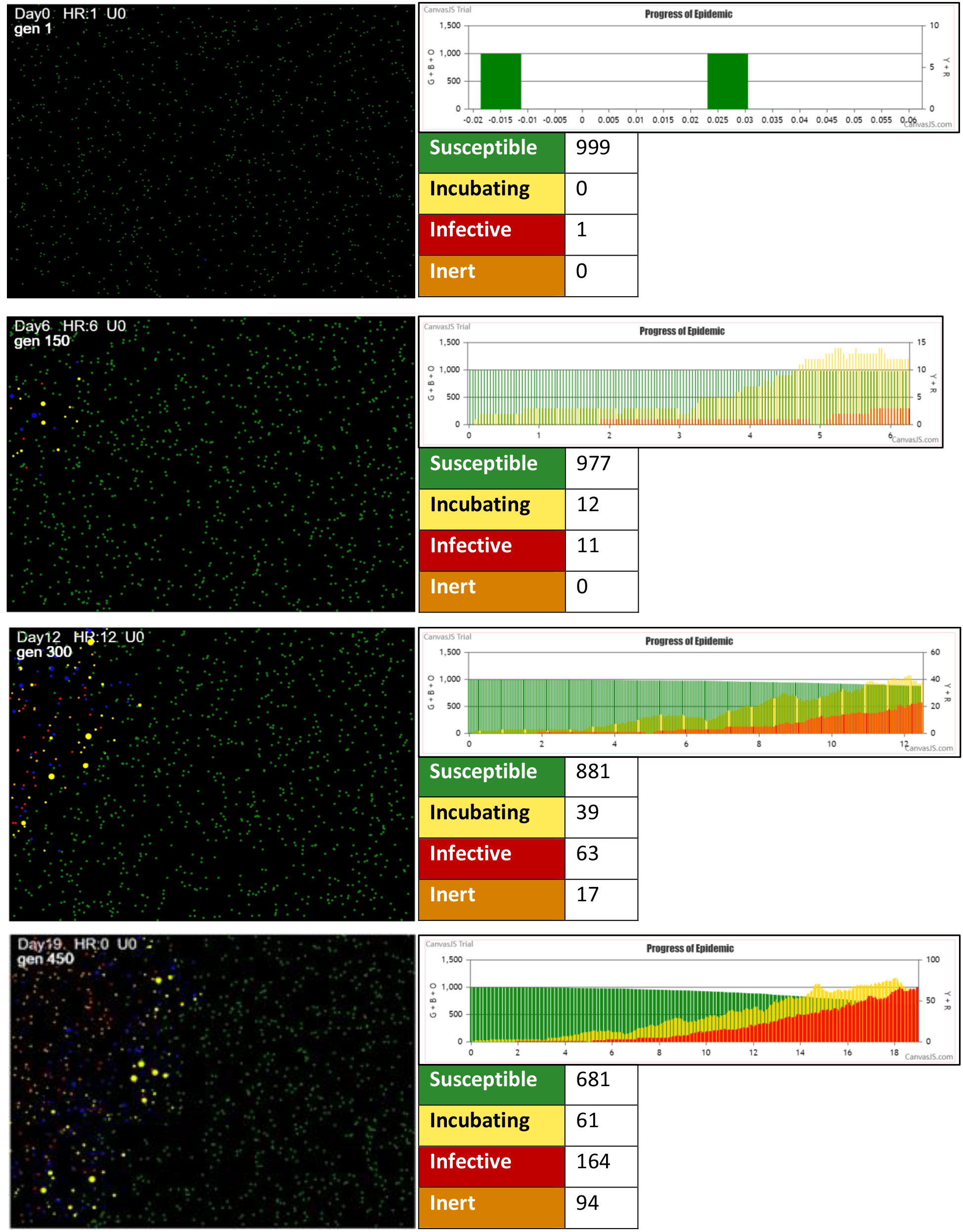

**WAVE DYNAMIC** - Parameters HzR = 2.1; mF = 0.95; RedDays = 8.2 **GENERATIONS 600 – 1050**

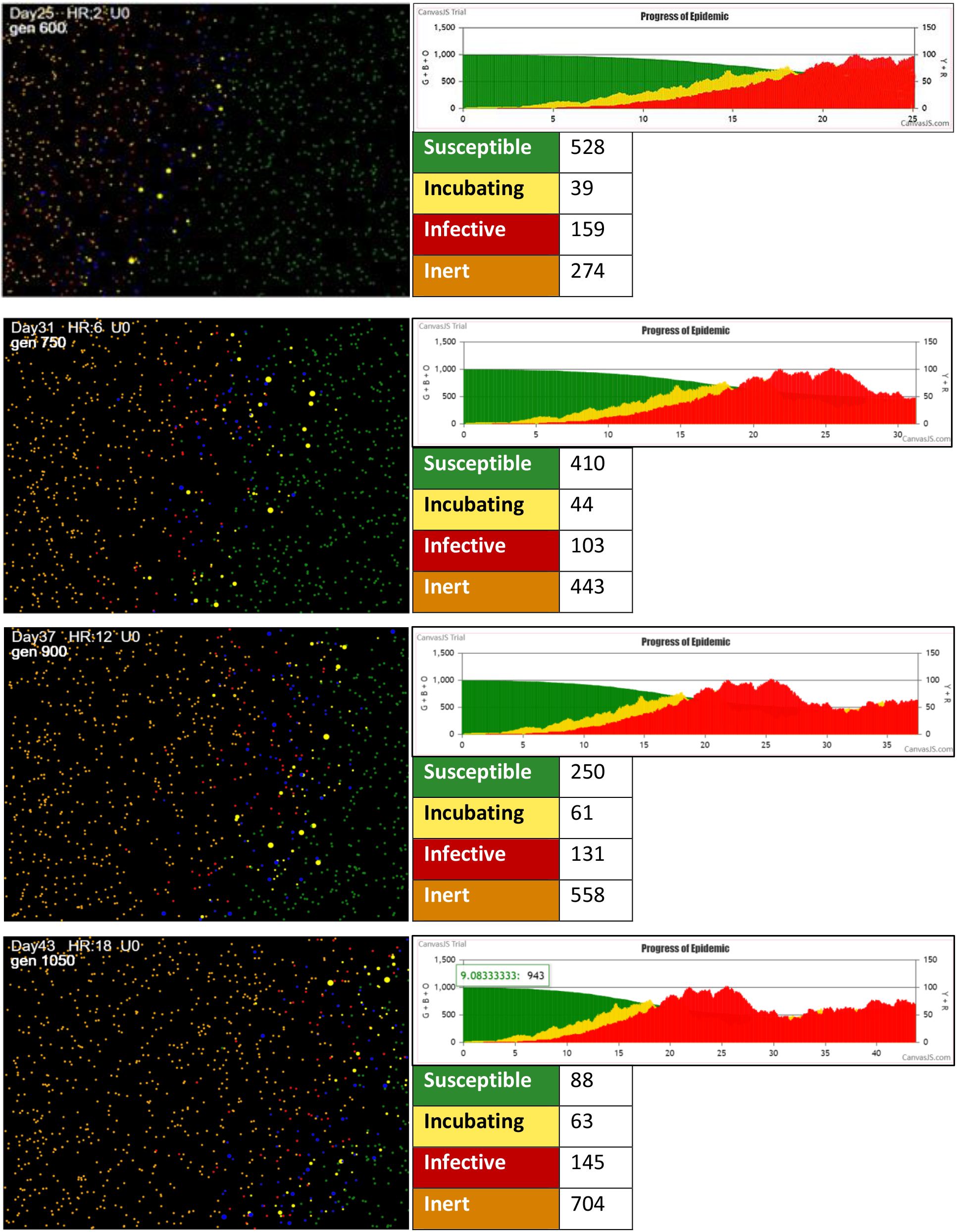

**WAVE DYNAMIC** - Parameters HzR = 2.1; mF = 0.95; RedDays = 8.2 **GENERATIONS 1200-TERMINATION**

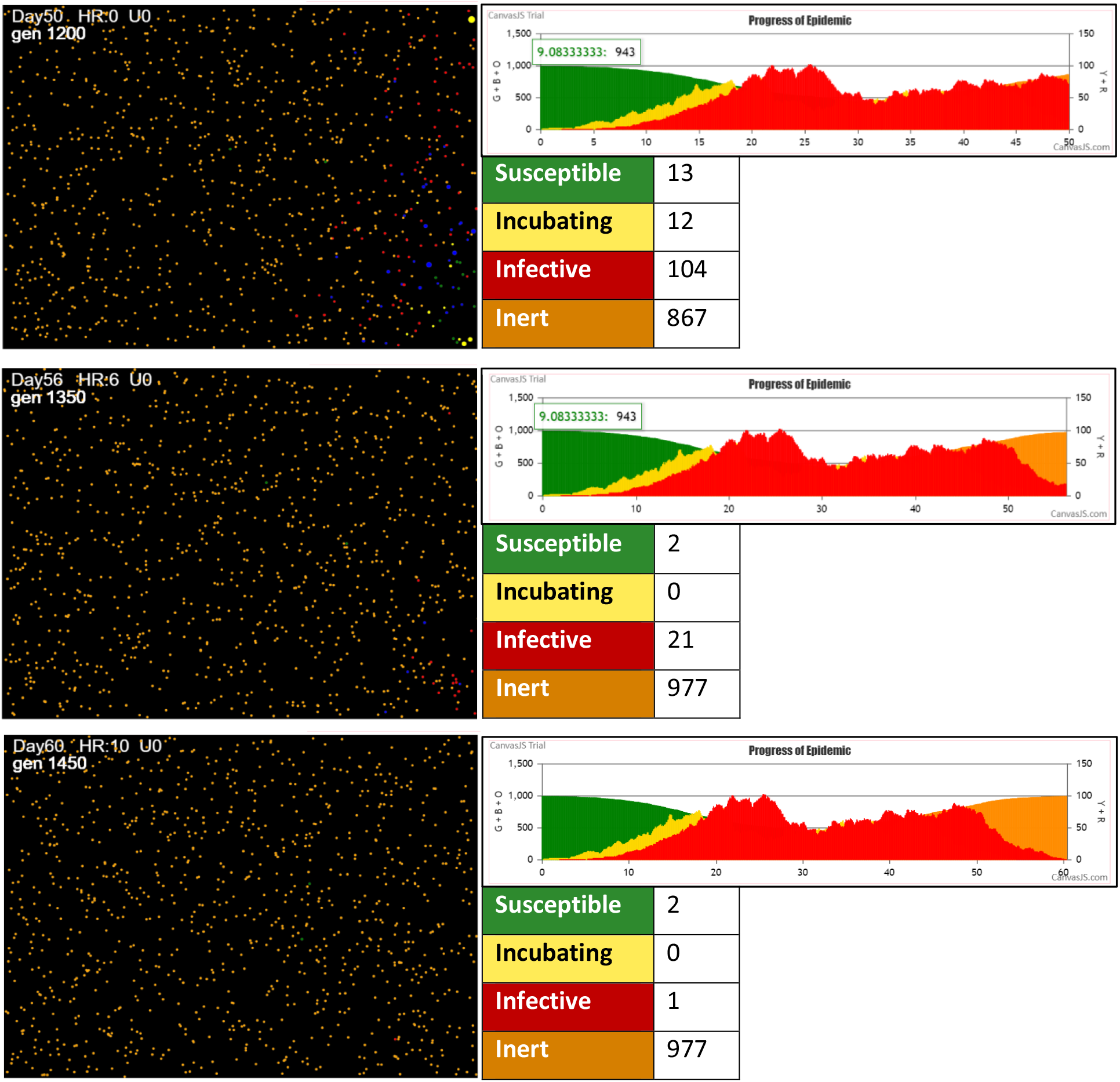

**Terminates at Generation 1454 with two remaining Susceptibles**.

## REFERENCES

1. Voit, E, H. Martens, S, Omholt (2015) 150 Years of the Mass Action Law. PLoS Comput Biol 11(1): e1004012. doi:10.1371/journal.pcbi.1004012. p.6

2. Chemistry LibreTexts. Mass Action Law. Last updated August 15, 2020. https://chem.libretexts.org/Bookshelves/Physical_and_Theoretical_Chemistry_Textbook_Maps/Supplemental_Modules_(Physical_and_Theoretical_Chemistry)/Equilibria/Chemical_Equilibria/Mass_Action_Law

3. Rate equation. https://en.wikipedia.org/wiki/Rate_equation#Second_order

4. OpenStax. Chemistry e2. 12.4 Integrated Rate Law. https://openstax.org/books/chemistry-2e/pages/12-4-integrated-rate-laws

5. Brauer, F. Compartmental Models in Epidemiology. 2008 In. In: Brauer F., van den Driessche P., Wu J. (eds) Mathematical Epidemiology. Lecture Notes in Mathematics, vol 1945. Springer, Berlin, Heidelberg. https://doi.org/10.1007/978-3-540-78911-6_7

6. Rebuli, N., N.G. Bean, J.V. Roos. Estimating the basic reproductive number during the early stages of an emerging epidemic. Theoretical population biology, 02/2018, Volume 119

7. Brauer, F. Chapter 4. An Introduction to Networks in Epidemic Modeling. In: Brauer F., van den Driessche P., Wu J. (eds) Mathematical Epidemiology. Lecture Notes in Mathematics, 2008, vol 1945. Springer, Berlin, Heidelberg

8. Ross J. On parameter estimation in population models III: time-inhomogeneous processes and observation error. Theor Popul Biol. 2012 Aug;82(1):1–17. doi: 10.1016/j.tpb.2012.03.001. Epub 2012 Mar 23. PMID: 22459805.

9. Moselle, K. A., & Ernie Chang (2020). CovidSIMVL – Agent-Based Modeling of Localized Transmission within a Heterogeneous Array of Locations – Motivation, Configuration and Calibration. medRxiv, (), 2020.11.01.20217943. Accessed January 06, 2021. https://doi.org/10.1101/2020.11.01.20217943.

10. Chang, E., Kenneth Moselle, Ashlin Richardson, A. (2020). CovidSIMVL --Transmission Trees, Superspreaders and Contact Tracing in Agent Based Models of Covid-19. medRxiv, (), 2020.12.21.20248673. Accessed January 06, 2021. https://doi.org/10.1101/2020.12.21.20248673.

11. He, X., E.H.Y. Lau, P. Wu. Temporal dynamics in viral shedding and transmissibility of COVID-19. Nat Med 26, 672–675 (2020). https://doi.org/10.1038/s41591-020-0869-5Xi He Transmission dynamics

12. Ibid.

13. Chang, Moselle & Richardson, op.cit.

14. Moselle & Chang (2020), op. cit.

15. Chang, E. & Kenneth Moselle. Agent-Based Simulation of Covid-19 Vaccination Policies in CovidSIMVL, January 21, 2021 medRxiv 2021.01.21.21250237; doi: https://doi.org/10.1101/2021.01.21.21250237

16. Pfizer-BioNTech COVID-19 Vaccine Emergency use Authorization Memorandum. November 20, 2020. https://www.fda.gov/media/144416/download

17. Moderna COVID-19 Vaccine VRBPAC Briefing Document December 17, 2020. https://www.fda.gov/media/144434/download

18. Chang & Moselle (2021), op. cit.

19. Bernal, J., Nick Andrews, Charlotte Gower, Chris Robertson, Julia Stowe, Elise Tessier, Ruth Simmons, Simon Cottrell, Richard Roberts, Mark O’Doherty, Kevin Brown, Claire Cameron, Diane Stockton, Jim McMenamin, Mary Ramsay., Early effectiveness of COVID-19 vaccination with BNT162b2 mRNA vaccine and ChAdOx1 adenovirus vector vaccine on symptomatic disease, hospitalisations and mortality in older adults in the UK: a test negative case control study https://khub.net/documents/135939561/430986542/Early+effectiveness+of+COVID+vaccines.pdf/ffd7161c-b255-8e88-c2dc-88979fc2cc1b?t=1614617945615 Posted Feb 19, 2021 Downloaded March 2, 2021

20. National Advisory Committee on Immunization (NACI): Statements and publications. Recommendations on the use of COVID-19 vaccines. January 12, 2021. https://www.canada.ca/en/public-health/services/immunization/national-advisory-committee-on-immunization-naci/recommendations-use-covid-19-vaccines.html#a2

