## Supplemental material for "Two Distinct Dynamic Process Models of COVID-19 Spread with Divergent Vaccination Outcomes"

CovidSIMVL

**Version:** 3.0

**Date:** April 7, 2021

#### 1 Introduction – good regulator theorem and epidemiological models

##### 1.1 Good regulator theorem

CovidSIMVL is an agent-based simulation modeling tool designed to capture key features of local contexts and associated transmission dynamics where contagion-based infection takes place. It represents an attempt to position modeling of dynamics governing viral transmission on a foundation that conforms to the basic requirements of Conant & Ashby's "good regulator theorem"[1], which may be viewed as a restatement or more explicitly formalized version of Ashby's Law of Requisite Variety[2]:

> *The first effect of this [good regulator] theorem is to change the status of model-making from optional to compulsory. As we said earlier, model-making has hitherto largely been suggested (for regulating complex dynamic systems) as a possibility: the theorem shows that, in a very wide class (specified in the proof of the theorem), success in regulation implies that a sufficiently similar model must have been built, whether it was done explicitly, or simply developed as the regulator was improved. Thus the would-be model-maker now has a rigorous theorem to justify his work.* (Conant & Ashby, 1970, p. 9)

As restated by Naughton[3]:

> *In colloquial terms Ashby's Law has come to be understood as a simple proposition: if a system is to be able to deal successfully with the diversity of challenges that its environment produces, then it needs to have a repertoire of responses which is (at least) as nuanced as the problems thrown up by the environment. So a viable system is one that can handle the variability of its environment. Or, as Ashby put it, only variety can absorb variety.*

In keeping with this law, CovidSIMVL seeks to model the dynamics that govern viral transmission at an explicitly contextualized person-to-person level where transmission of viruses such as SARS-CoV-2 takes place. The overarching objective is to supply models that can be applied to decision-making (i.e., regulating pandemic response) that relates to entities far smaller than the populations-at-large to which typical equation-based models refer.

##### 1.2 Why agent-based modeling?

A sample scenario will make the potential scope and relevance of a tool like CovidSIMVL more clear. Consider a health service region that is responsible for the vast majority of secondary and tertiary health services. To contend with the possibility of healthcare associated transmission in the midst of a pandemic, decisions need to be made regarding services to keep open despite possibly rising rates of infection in the community *vs* services to close down temporarily. While modeling is probably not necessary to consider questions around services such as Emergency Departments or Med/Surg Acute Care units and a diverse array of collateral clinical supports (e.g., Medical Imaging, laboratory), there remain many other services to consider, for example, a broad array of ambulatory services, medical imaging not associated with acute care admissions, or various community-based services including case management, home support, a diverse array of services for homeless persons possibly contending with various combinations of severe psychiatric illness, chronic-relapsing addictions, rehabilitation services, etc.

Forecasts generated from equation-based compartmental models of epidemic spread may help to calibrate decision-making when those decisions are to be applied to the population at large, based on best-guesses about future base rates of infection in the population at large. Or restated in terms of the good regulator theorem, those forecasting models may enable state/provincial or federal public health agencies to regulate pan-societal response to pandemic spread. However, those models are challenged to drill down to the granular level at which distinct services are delivered. In the case of a health region such as Island Health in British Columbia, that granularity is reflected in over 1970+ service units arising from the location build in the service system's full cross-continuum deployment of an electronic health record.[4]

Further, the "population" to which the equation-based models relate is not homogeneous with respect to patterns of service utilization and risks associated with service access. Meta-population models or "patch-models" may enable generation of slightly more granular versions of population-level compartmental models, but those approaches are limited as to how far they can "reach" towards the real-world contexts in which transmission takes place.[5] If the concern, for example, is risk for inadvertent cross-over transmission to or from at-risk populations *via* access to health services, then models must be able to treat populations at a more granular level and relate those smaller units to contexts, and then go from people in contexts to risk.

##### 1.3 CovidSIMVL – from Context to Dynamics, from Dynamics to Events

This document provides an overview of CovidSIMVL, which is an agent-based modeling tool used to simulate spread of contagion-based viral infections such as SARS-CoV-2. The document provides conceptual clarity around the constructs that guide the configuration of CovidSIMVL to reflect dynamics of local "real-world" scenarios. By enabling the conduct of virtual clinical trials in which variations are introduced systematically into a range of potential determinants or risk factors (e.g., behaviour within and across local contexts) and interventions (e.g., vaccination schedules), the intent is to supply a means for projecting potential impacts of various contemplated localized options for managing pandemic spread.

CovidSIMVL is an open-source public domain system freely available under the GNU Open License Framework, and can be found at www.github.com/ecsendmail/MultiverseContagion. It is written in Javascript and runs in most modern browsers such as Chrome, Edge, Safari and Firefox. See ***Figure 1*** for an illustration of CovidSIMVL simulation of an epidemic taking place in a single bounded space ("Universe").

#### 2 Objectives of CovidSIMVL

CovidSIMVL has been architected and implemented to achieve two overarching objectives. The first is to enable simulation of localized processes of contagion-based viral transmission (transmission dynamics).

The second is to explore the likely impacts of various risk factors and protections, by looking at their proportionate contribution to measured outcomes (virtual clinical trials).

***Figure 1.*** *CovidSIMVL – Single Universe Simulation.*

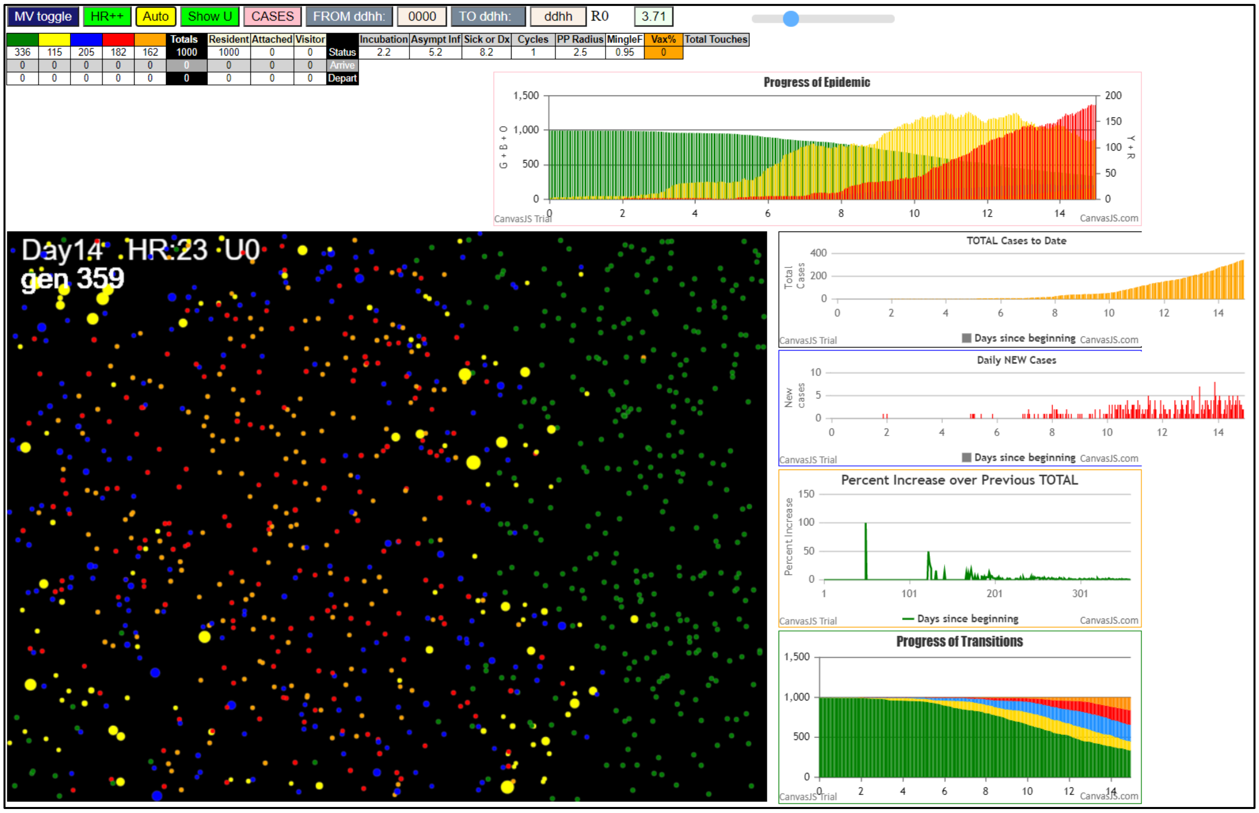


#### 2.1 Model transmission at a level of concreteness, granularity, and heterogeneity to reflect real-world contexts and dynamics governing transmission.

Contagion-based infection is anchored in processes that create a clinically impactful relationship between viral agents and persons. In the case of SARS-CoV-2 this is the epitome of a person-to-person relationship, with infectivity rapidly dropping off as people are separated by distance or time. As such, the foundational contextual frame for any CovidSIMVL simulation is a local context (a CovidSIMVL "Universe") in which people with transmission-relevant properties (human "agents") engage with one another.

#### 2.2 Enable the conduct of virtual clinical trials.

Parameters in CovidSIMVL are variables that can be set to reflect differences in those factors that impact on transmission. For example, different levels of presumed or hypothesized inherent infectivity can be embodied in parameters in the model that do not vary as a function of the behaviour of the agents. By contrast, impacts associated directly with the behaviour of individuals can be captured *via* parameters that translate into movement of agents that create the possibility for infection to occur by altering their spatial separation. Other features of CovidSIMVL can be altered to change characteristics of roles, or movement of agents across different contexts (e.g., school, home, place of work). As well, rules that govern the probability of transmission or duration of infectivity can be altered to reflect expected rates of protection associated with vaccination or the institution of other protections.

By holding some of these parameters constant while varying others, CovidSIMVL virtual clinical trials can be conducted to evaluate the impacts of factors and, depending on the design, to evaluate their interactions.

Because CovidSIMVL is a computerized simulation modeling tool, virtual trials can be conducted when various factors such as time or logistics or ethics preclude the possibility of running comparable trials in real life.

#### 2.3 Modeling causes *vs* predicting effects (events)

Equation-based compartmental models in epidemiology take events as their inputs (e.g., daily confirmed new cases or hospitalizations or deaths) and output systems of equations, with values assigned to parameters in order to close the distance between predicted values and measured values.[6]

Agent-based models take encoded specifications of transmission dynamics as "inputs" and generate events (e.g., infections over time) as outputs. These outputs can then be taken as inputs for equation-based modeling or they can be used to generate other mathematical/statistical summaries of the sequences/sets of events that emerged in the context of the agent-based simulations.

Equation-based models are often used to forecast future states of affairs. For example, if they are based on population-level data, and they make certain key assumptions (e.g., homogeneous spread, see ***Sec. 4.2***, below) they can be used to project impacts of various contemplated risk mitigation strategies for the population as a whole. This can be useful if "pan-societal" policies (applied to all members of the population) are under consideration, in order to mitigate risk for pandemic spread, e.g., "all members of the public must remain at home except for essential health service providers".

CovidSIMVL is not designed to predict the future, e.g., expected rates for events such as new cases or hospital admission in a real-world geographically bounded locale, conditional upon certain steps being taken to impact on those rates. It is designed to model the dynamics that give rise to events, such as new cases of infection. These events can then be analyzed and modeled to evaluate the impact of those factors that govern the dynamics out of which the events have emerged. This may be useful when localized measures are contemplated in order to maximize the welfare of the public at large while reducing the scope of protective measures taken to achieve that larger good.

There is no expectation that CovidSIMVL could scale out to make predictions about real-world rates for events over time if it had access to a sufficiently powerful computing facility. Any given CovidSIMVL simulation characterizes the progress of an infection as it navigates across an epigenetic terrain in which phenotypical characteristics of the SARS-CoV0-2 agent are heavily impacted by behaviour, heterogeneity in key spatial and temporal features, and stochasticity – see ***Sec 4***. There is no expectation that the model could predict the future in real-world situations unless it had access to "meta" information about those factors that determine human behaviour, and until those parameters that determine human behaviour were encoded and reflected in rules in CovidSIMVL, which is not currently the case. Even with access to this full body of information, essential stochastic features of viral transmission would continue to play out in possibly marked variation in spread.

### 3 Functions Performed by CovidSIMV

CovidSIMVL is designed to perform three essential functions, in order to achieve the objective of simulating localized contagion-based viral spread and highlighting the impacts of factors in the model that determine spread:

#### 3.1 Reflect inherent properties of the viral agent.

CovidSIMVL models essential pathophysiological properties of contagion-based viral transmission, e.g., rising/falling viral load and associated changes in infectivity, or vaccine-produced changes in risk for becoming infected or infecting others.

#### 3.2 Capture the influence of a potentially heterogeneous array of contextual and/or behavioural factors on transmission

CovidSIMVL rules embody those factors that interact with inherent properties of a viral agent to produce viral transmission, or protect against transmission. Through this interaction of intrinsic (to the virus) and extrinsic factors (e.g., density of agents or movement within a setting), local realism is incorporating into modeled processes of contagion-based viral infection.

#### 3.3 Supply summary metrics and companion visualizations

To translate CovidSIMVL-generated events into contents (metrics, visualizations) that can be compared, characterized and communicated, CovidSIMVL layers models onto outputs in a manner that is analogous to the creation of equation-based SEIR models from daily counts of confirmed new infections.

1. Some of these models are expressed in terms of quantities, such as time between new infections (CovidSIMVL "theta"), or statistical properties of events, e.g., changes in "velocity" of transmission. See for example ***Figure 1***, top-middle, a reproduction number (R0) of 3.71, representing the average number of secondary infections produced by infective agents at the particular point in the simulated outbreak depicted in the screen capture.
2. These models may also reflect characteristics of the transmission dynamics over time, e.g., the topological structure of chains of transmission, or rooted tree structures formed by chains.

### 4 The CovidSIMVL Viral Transmission "World-View" – Heterogeneous Dynamics, Stochasticity

#### 4.1 Localized transmission

CovidSIMVL concerns itself with inherently localized contagion-based transmission of disease – from one agent to another, embodying mechanisms such as droplet transmission in the rules that determine whether movement of agents in a space produces transmission. As such, CovidSIMVL is suited to the task of modeling contagion-based transmission of viral agents such as SARS-CoV-2.

CovidSIMVL is not designed to model vector-borne diseases such as malaria or dengue fever (carried by mosquitos) or food- or water-borne diseases carried by biological agents (e.g., E-coli; cholera) or by non-communicable diseases that result from exposure to toxic environments (e.g., radiation, lead poisoning from pipes/drinking water).

#### 4.2 Spread *via* Context-Bound Heterogeneous Contact vs Context-Free Mass Action

Equation-based epidemic models typically incorporate some version of the "law of mass action" which is embodied in models of infectious disease spread "…in which all individuals in a population make contact at an identical rate and have identical probabilities of disease transmission to those contacts per unit of time."[7] This assumption is counterfactual but not necessarily consequential if the intent is to make predictions based on datasets where there are large numbers of persons in each of the Susceptible/ Incubating/Infective/Recovered groups. In this case, differences in contact patterns will average out and the prediction models may remain robust.

However, if the intent is to understand the processes that govern spread, then it does not make sense to work with models that are built on a foundation of known counterfactual assumptions about those processes.  Where the intent is to understand dynamics – which is the principal objective of CovidSIMVL – then the models must enable real-world heterogeneity to be injected into the models.[8] As well, if the number of infectives is small, which is always the case in simulations that start with a single infective agent located in an array of susceptible parties, then the model must incorporate stochastic effects.[9]

See ***Figure 2*** for a CovidSIMVL simulation that conforms to a "mass action incidence" model. As in all CovidSIMVL visualization, Green = Susceptibles; Yellow = Incubating; Blue = Asymptomatic Infectives; Red = Sympatomatic Infectives. In ***Figure 2***, the parameter that reflects inherent infectivity have been set to a moderately low value (see ***Sec. 7.2.1***, below) while the parameter that reflects movement and mingling have been set to a high value (see ***Sec. 7.2.2***).  In this simulation, agents in different states are well dispersed throughout the space by Generation 255 – about 11 days.

By contrast, see ***Figure 3*** for a CovidSIMVL simulation that has been configured to illustrate spatial heterogeneous spread. In this visualization, the parameter that reflects infectivity has been set to a high value, while the parameter that reflects movement and mingling has been set to a low value.

In this figure***,*** we see the spread, which started with a single Infective agent in the upper right-hand corner, advancing from the upper right diagonally across a dispersed array of Susceptibles.  More specifically, we see a spatially (and temporally) advancing wave of Incubating agents, followed by a wave of Asymptomatic Infectives (Blues), followed by a was of Symptomatic Infectives (Yellows).

The rough, by no means complete or clean separation of agents into coloured bands reflects stochasticity built into the transmission dynamics in CovidSIMVL.

To summarize: What we see are two different dynamics based on configuration of parameters that embody real-world characteristics of contagion-based infectious spread, namely, degree of infectivity and movement of agents in a finite space. Further to this point: see Chang & Moselle (2021a).[10] In addition to supplying more detail on distinctive features of these two dynamics, together with metrics that reflect those differences, that paper also reports on simulated vaccination trials, to illustrate different outcomes associated with different dynamics.

***Figure 2.*** *Simulation of "mass action" spread  of infection – a "particle" dynamic simulated in CovidSIMVL*

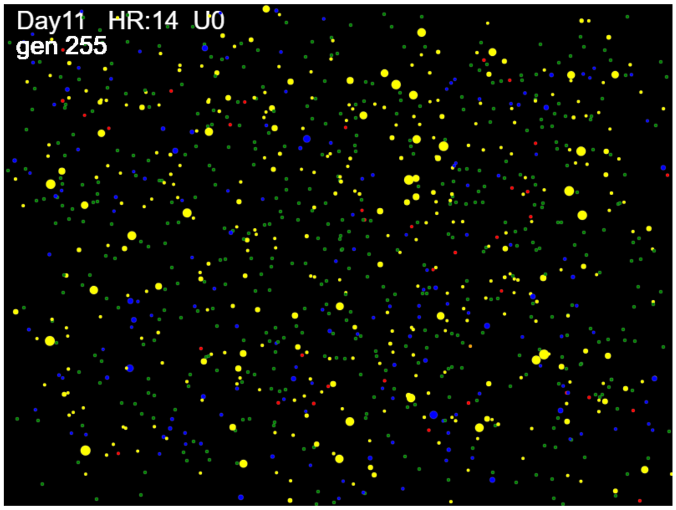

***Figure 3.*** *Simulation of a "wave" dynamic of infection spread in CovidSIMVL*

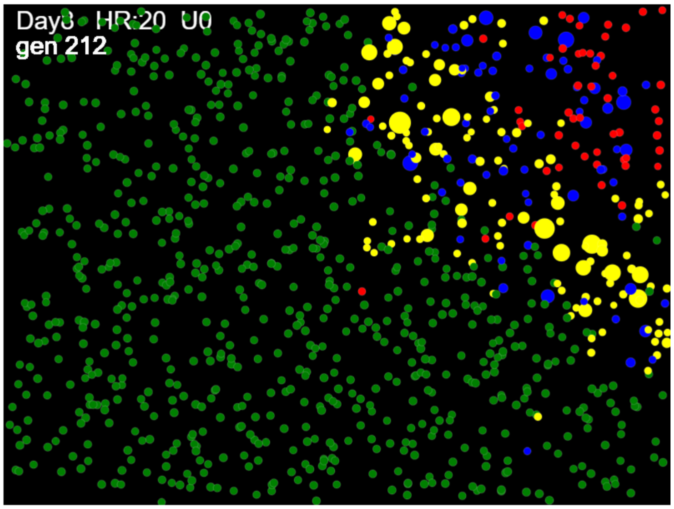


#### 4.3 Heterogeneity and Stochasticity in CovidSIMVL Configuration and Dynamics

##### 4.3.1 Heterogeneity

###### *4.3.1.1 Factor 1 – Inherent characteristics of agents – their "states".*

In the case of contagion-based infection, agents maybe Susceptible, Exposed and incubating but not infectious, Infectious, or removed in some fashion from the population, e.g., *via* recovery from illness, death, or quarantine ("Inert" in CovidSIMVL; often referred to with the letter "R" or "Removed" in compartmental epidemiological models). These partitions map directly onto the compartments in classic SEIR equation-based models.

###### *4.3.1.2 Factor 2 - Behavioural characteristics of agents; geospatial partitioning of the "real world".*

These reflect the person and the physical spaces or contexts or roles in which they are operate. For example, an employed adult will move and interact differently at home, in a large food store, in a gym, or in a place of work. A realistic depiction of this agent would need to reflect both the changes in roles and the characteristics of the contexts in which the agent is acting in a role-determined fashion.

###### *4.3.1.3 Factor 3- "Other" or "hybrid",*

These are factors which may reflect an interaction among Factors #1 and #2 directly above. Age would be a good example. People of different ages may differ in their physiological response to an infectious agent, and they may also spend time in different locations and move differently in those locations.

Factor #3 can turn out to be methodologically "thorny" for epidemiological models. Age, for example, may turn out to be predictive of some characteristic of viral transmission. However, we may ask whether measured effects associated with age a reflection of the inherent properties of the virus or are they a reflection of differences in behaviour between different age cohorts? Equation-based models may attempt to tease apart these factors by segmenting populations and generating meta-population models or "patch" models.[11] This approach has been proven out when the number of patches is small. However, this approach can run into computation or other issues, e.g., relating to sample size, when the number of factors or patches increases to reflect the local "topography" of heterogeneous transmission in real-world contexts.[12] There may also be challenges when these factors to be evaluated are not directly measured but are instead grafted on to the study population from some other source.

By contrast, because Factor #1 and #2 are encoded as essential features of every agent within every CovidSIMVL simulation, these components can be controlled and their effects can be measured directly in the simulation. Thus, the tool is well suited to studying the potential for either or both sets of factors to contribute to the dynamics of transmission, to the topological characteristics of chains or trees that graphically depict those dynamics, and to the metrics that summarize those dynamics and topological features.

##### 4.3.2 Stochasicity

In any given CovidSIMVL simulation, values are assigned to rules which determine when transmission occurs. These rules are not strictly deterministic. If they were, you could know exactly how many agents would become infected for a given trial if you knew the initial values for all parameters and you knew the initial conditions.

In CovidSIMVL, one set of parameters could result in a simulated epidemic in which every agent becomes infected within 1000 iterations (Generations). The same parameters could result in an outbreak that self-terminates after 350 iterations, because all infectious agents by the 350$^{th}$ iteration of the trial had become "Inert" (non-infectious) before they had transmitted to any other agents.

This reflects the real world, where various factors (e.g., persons masked vs unmasked; physical distance; wind or air movement, etc.) will determine whether a person with a sufficiently high viral load will transmit to another person.

It is important not to equate "stochasticity" with "randomness. Stochastic agent based models of processes make use of random probability distributions (e.g., Gaussian normal curve; negative binomial distributions) to govern features of the behaviour of agents. For example, in CovidSIMVL, a value is drawn randomly from a standard Pareto distribution to introduce some degree of variability in the movement of agents within a physical space. This will have *some* impact on what proportion of a population of Susceptible agents in CovidSIMVL end up being infected. However, agents are not selected at random to become infected.

#### 4.4 Basic Reproduction Number (RO), Effective Reproduction Number (R) in CovidSIMVL

The "reproduction number" or "basic reproduction number" (R0) is an aggregate figure, intended to reflect the expected number of secondary infections arising from any given index case within a completely susceptible population. Within a mass action incidence model, it would be expected to provide the best statistical estimate of the number of secondary infection events likely for any given member of a given population.

Caution must be exercised in using quoted R0 values to guide decision-making or policies:

> *The basic reproduction number (R0), also called the basic reproduction ratio or rate or the basic reproductive rate, is an epidemiologic metric used to describe the contagiousness or transmissibility of infectious agents. R0 is affected by numerous biological, sociobehavioral, and environmental factors that govern pathogen transmission and, therefore, is usually estimated with various types of complex mathematical models, which make R0 easily misrepresented, misinterpreted, and misapplied. R0 is not a biological constant for a pathogen, a rate over time, or a measure of disease severity, and R0 cannot be modified through vaccination campaigns. R0 is rarely measured directly, and modeled R0 values are dependent on model structures and assumptions. Some R0 values reported in the scientific literature are likely obsolete. R0 must be*

*estimated, reported, and applied with great caution because this basic metric is far from simple.*[13]

Note the statement "R0 is rarely measured directly." This is true for equation-based compartmental models, where it is estimated from confirmed case counts over time for a given dataset. It is also true for many agent-based models, where R0 is set at a fixed value when the simulation is initialized.

It is not true for CovidSIMVL. In CovidSIMVL, transmission is a joint function of the inherent properties of the viral agent and the spatio-temporal dynamics of the agents' movements within one or more spaces. Reproduction of the virus *via* transmission to a new host can take place, subject to stochastic variation in both viral shedding and in agent movement. If a susceptible agent is sufficiently close to an infectious agent in CovidSIMVL, transmission can occur. R0 is derived by counting within agents and averaging across agents. It can be measured at any point in time (i.e., Generation) within a simulation.

#### 4.5 Vaccination

The one important exception to the operational rule that transmissions rates are not pre-set in CovidSIMVL relates to vaccination effects. In this agent-based modeling tool, vaccination success rates are treated as fixed cut-offs that are used to evaluate whether proximity of an infective and susceptible agent gives rise to transmission. Each agent in a simulation is assigned a random number ranging from 0 to 1. When a proximity threshold is exceeded, the randomly assigned number is evaluated against a threshold set by a vaccination success rate, e.g., 75%. Infection will occur only if the susceptible agent's randomly assigned value exceeds .75. Thus, the fixed vaccination success rate does not determine exactly which vaccinated agent does/does not become infected when contacted by an Infective. In other words, vaccination effects are layered onto the spatio-temporal dynamics of CovidSIMVL and are still subject to stochastic effects.

### 5 Calibration *vs* Validation of Simulation Models

CovidSIMVL parameters are calibrated by determining the ranges that they can take in order to reproduce sets of events (e.g., daily counts) that track to curves associated with standard metrics such as R0. See Moselle & Chang (2020)[14] for details. The dynamics governing transmission may be regarded as "validated" to the extent that the parameters can by configured to reproduce ***any*** set of outcomes in any real-world contexts where the dynamics are thought to be governed by factors that are embodied in those CovidSIMVL parameters.

### 6 CovidSIMVL – Spatio-Temporal Architecture – Single Universes; the CovidSIMVL Multiverse

#### 6.1 Viral transmission within a spatio-temporal frame of reference.

In CovidSIMVL, viral transmission occurs within three temporal frames of reference and within one or more physical spaces.

##### 6.1.1 Temporal frames of reference

###### *6.1.1.1 Trials*

The most fundamental unit of duration in CovidSIMVL is a trial. A trial proceeds through a set of iterations. At each iteration, characteristics of agents may change, e.g., an agent might move to a point of close proximity to an infective agent, or protections associated with a vaccination may lapse if a second dose is not administered, or an agent may move from one Universe (e.g., home) to another (e.g., place of work).

A trial concludes when specified conditions are met. These conditions generally fall into two classes:

1. Self-extinction - there are no more Infective agents, so additional iterations will not give rise to any new infections.
2. Exceeding a threshold value - infective agents remain but a specified benchmark or cut-off is reached. For example, a trial may conclude when 50% of the original Susceptible agents have become infected.

Measurements may be taken when a trial reaches the point of self-extinction. As well, measurements may be taken when various thresholds are reached or intervals are spanned, e.g., value of the CovidSIMVL theta metric (time between transmissions) for an interval extending from 10% to 19% of the original Susceptibles have become infected.

###### *6.1.1.2 Generations – Basic Units of Time*

A single CovidSIMVL trial consists of a series of events that emerge over the course of iterations, subject to rules as detailed in ***Sec. 7***. At each Generation (i.e., iteration) in a trial, the full set of rules are applied to every agent. Some of these rules are subject to stochastic variation as described in that section.

Time-dependent events or time-denominated metrics are all keyed to Generations, and no changes of state or events occur until the Generations counter is incremented.

Generations can be treated as a purely scalar quantity in CovidSIMVL. However, Schedules (see ***Sect 7.3***, below) provide a "real-world-clock" anchoring point for Generations. These Schedules, which describe the movement of agents within and between locales, are designed to reflect patterns of movement over the course of a real-world 24-hour day. By breaking the day into 24 units and iterating CovidSIMVL 24 times over the course of a given day, Generations come to acquire a unit, namely, one hour.

###### *6.1.1.3 Temporal Dynamics of Viral Transmission*

Within boundary limits set by stochastic variation, agents in CovidSIMVL who have been exposed to a viral agent remain in different states for set periods of time. These states are:

- Susceptible (but not contacted) – "Greens" in CovidSIMVL – agents remain in a Susceptible state indefinitely, so long as they are not located at any Generation in close proximity to an infective agent
- Incubating (but not infective) – "Yellow" in CovidSIMVL visualizations
- Infective but not symptomatic – "Blues" in CovidSIMVL visualizations
- Infective and symptomatic – "Reds" in CovidSIMVL visualizations
- Inert – "Oranges" in CovidSIMVL

###### *6.1.1.4 Temporal durations for parameters that reflect infectivity.*

Temporal dynamics in CovidSIMVL are derived from the model of He and associates.[15] This model (see ***Figure 4***) sets working reference ranges for viral loads and days after infection for incubation, pre-symptomatic transmission, first appearance of symptoms, and viral load tapering to below measurement thresholds in mild cases. In CovidSIMVL, these durations are translated into Generations.

***Figure 4**. Configuring Primary Rules – Temporal Dynamics in Viral Shedding*

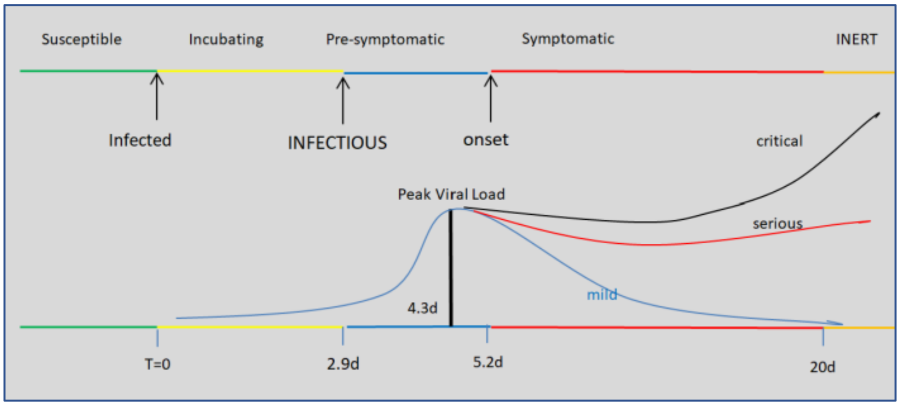


###### *6.1.1.5 Scheduled movements of agents between Universes in CovidSIMVL Multiverse Simulations*
In every Multiverse simulation, every agent is associated with a schedule of movements between Universes. Any given agent's risks for transmission may be changed as they cross over from one location to another by altering the way the agent moves within a given location. In other words, these characteristics that govern risk are bound to agents, but may vary systematically by location.

###### *6.1.1.6 Vaccination timings*
Simulations involving vaccinations require values on three parameters relating to timing:

- Elapsed time between vaccine administration and onset of protection
- Timing between dose for two dose regimens
- For vaccines that specify two doses to achieve full or sustained protection - duration of protection if only the first of a two-dose regiment is administered.

##### 6.1.2 Spatial frames of reference

There are two spatial frames of reference in CovidSIMVL – Universes, and Multiverses, which consist of functionally interconnected Universes.

See ***Figures 1- 3,*** above, for screen shots of single Universe simulations in CovidSIMVL.

A Universe in CovidSIMVL has finite dimensions. These dimensions determine the density of agents within a given CovidSIMVL Universe. Because transmission is a function of proximity, density will impact on the rate of transmission.

A Multiverse consists of a collection of Universes. See ***Figure 3***, below. Each agent has a schedule that determines their location in any given Universe over the course of Generations in a CovidSIMVL trial.

### 7 Setting Parameters in CovidSIMVL

Abstracted from Chang, Richardson & Moselle (2020):[16]

CovidSIMVL enables model calibration by employing rules organized in a three-fold hierarchy to support the injection of biological, behavioural and local spatially-contextual heterogeneity that we deem to be logically necessary to support a diversity of realistic model scenarios, as per broad methodological principles set by the “Law of Requisite Variety” or the closely-related “good regulator theorem”.

***Figure 2**. CovidSIMVL – Multiverse Simulation*

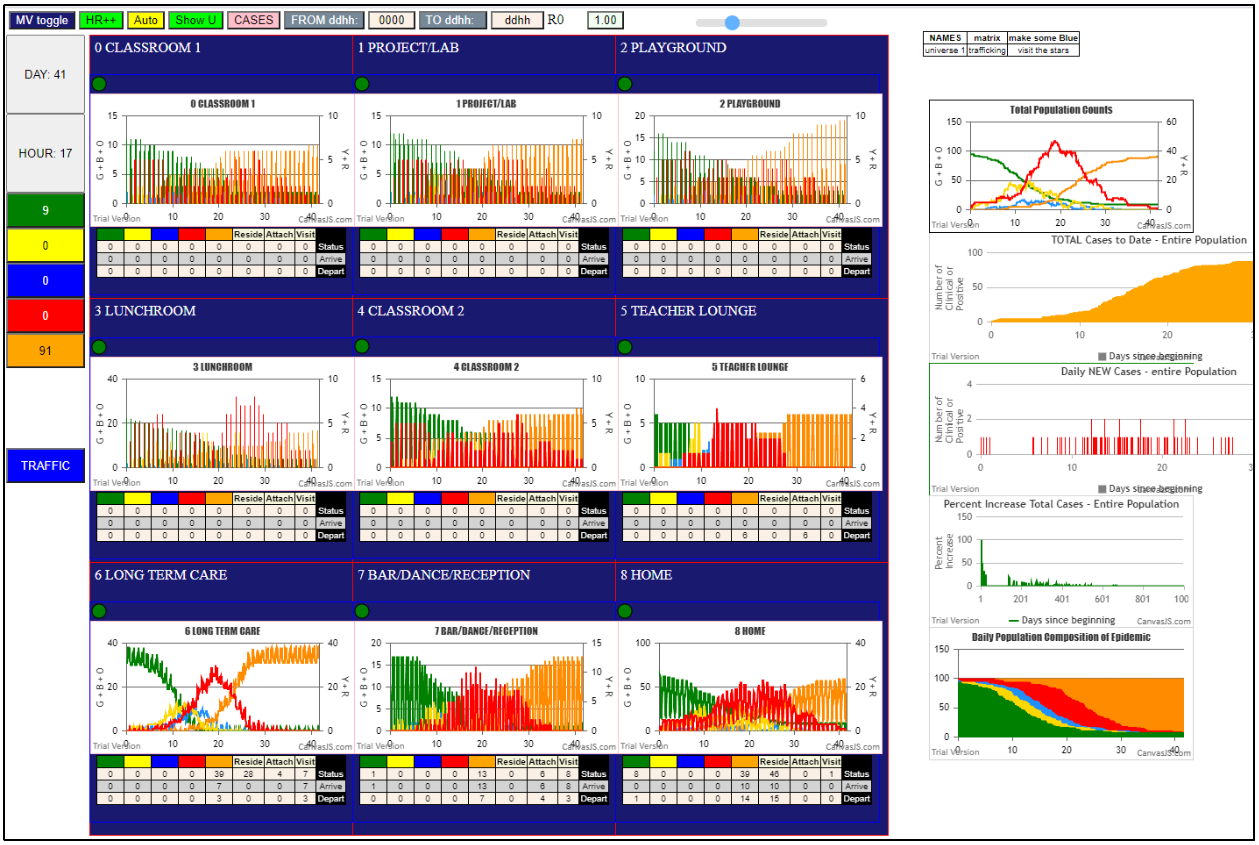


#### 7.1 Primary Rules

These are intended to embody physiologically determined viral spread features e.g., usual incubation period or duration of infectivity. Because viral transmission reflects processes of epigenesis, in which genomic characteristics interact with environmental factors to determine phenotypic characteristics, these primary rules may become the functional "repository" of variations in either genomically-determined characteristics, or environmentally-determined characteristics.

For example, in CovidSIMVL, the variable "RedDays" can be set at the outset of a trial or midway through a simulation to reflect the number of days that a symptomatic agent can infect others. In the real word, this is determined in part by the properties of the virus and in part by the behaviour of the person, as determined by the person or context. If new data became available suggesting that this period of time could be longer or shorter than was previously believed to be the case, RedDays could be increased or reduced.

Along those same lines, if proactive testing were put in place such that symptomatic people transitioned into an "Inert" state more rapidly, then RedDays could be shortened accordingly.

#### 7.2 Secondary Rules

These are rules or parameters that determine if/when/where a susceptible person becomes infected or when/where infectious transmission occurs. For example, probability of transmission varies with proximity, which is modulated in CovidSIMVL according to the HazardRadius and MingleFactor parameters:

##### 7.2.1 HazardRadius (HzR)

This parameter functions as a secondary rule to determine transmission risk within a microcosmic spatial reference frame (a single CovidSIMVL Universe). Specifically, HazardRadius determines the likelihood of transmission between agents when they are located within specified distances from one another in a given simulation trial Generation. Protections such as masking would be reflected in lower values on HazardRadius.

##### 7.2.2 MingleFactor (mF)

This parameter incorporates movements of agents in a delimited Universe. Subject to stochastic variation according to a Pareto "random walk" distribution, the MingleFactor parameter, over the course of iterations, determines how many susceptible or infectious agents fall within a critical radius where transmission can take place.

#### 7.3 Tertiary Rules – CovidSIMVL Schedules

These Schedules set rules/parameters that prescribe patterned movements of agents within and between local contexts. Specifically, CovidSIMVL Schedules describe movements of agents in local Universes. These schedules are immediate determinants of intra-Universe interactions. Secondary rules may be assigned to agents within specific universes, and these may change as the schedule prescribes movement to other Universes – an agent may behave in a manner that creates a higher level of infection risk in a pub than in a place of work. They also function as determinants of cross-over transmission between Universes.

Schedules include a specification of local contexts (e.g., School, Home, Long-Term Care Facility); social roles (e.g., Student, Teacher, Spouse, Long-Term Care Resident), and family structures (e.g., single persons; two parents and two children and one grandparent).

For details on configuration of schedules, see Chang & Moselle (2021b).[17]

REFERENCES

1 Conant, R.C. and Ashby, W.R. (1970), Every good regulator of a system must be a model of that system, Int. J. Systems Sci., 1970, vol 1, No 2, pp. 89–97

2 Ashby, W.R. (1956), An Introduction to Cybernetics, Chapman & Hall, 1956, ISBN 0-416-68300-2

3 Naughton,. J. 2017: What scientific term or concept ought to be more widely known. Edge. https://www.edge.org/response-detail/27150. Downloaded March 21, 2021

4 Moselle, K. & Koval, A. Clinical Context Coding Scheme. March 28, 2019. Available on request from

5 Ball, F., Britton, T., House, T., Isham, V., Mollison, D., Pellis, L., and Scalia Tomba, G. (2015) Seven challenges for metapopulation models of epidemics, including households models Epidemics Volume 10, March 2015, Pages 63-67 https://doi.org/10.1016/j.epidem.2014.08.001

6 Brauer, F. (2008) Compartmental Models in Epidemiology. 2008 In In: Brauer F., van den Driessche P., Wu J. (eds) Mathematical Epidemiology. Lecture Notes in Mathematics, vol 1945. Springer, Berlin, Heidelberg. https://link.springer.com/chapter/10.1007/978-3-540-78911-6_2

7 Volz, E. and Meyers L. (2007) Susceptible-infected-recovered epidemics in dynamic contact networks [published correction appears in Proc Biol Sci. 2008 Dec 22;275(1653):2898]. *Proc Biol Sci*. 2007;274(1628):2925-2933. doi:10.1098/rspb.2007.1159

8 *Ibid*.

9 Brauer, *op. cit*., p. 45

10 Chang E. and Moselle, K. (2021a) Two dynamic process models of Covid-19 with divergent vaccination outcomes. Unpublished manuscript, March 21, 2021. Available on request from or medRxiv.

11 van den Driessche P. (2008) Spatial Structure: Patch Models. In: Brauer F., van den Driessche P., Wu J. (eds) Mathematical Epidemiology. Lecture Notes in Mathematics, vol 1945. Springer, Berlin, Heidelberg. https://doi.org/10.1007/978-3-540-78911-6_7

12 Ball *et al, op. cit.*

13 Delamater PL, Street EJ, Leslie TF, Yang YT, Jacobsen KH. (2019) Complexity of the Basic Reproduction Number ($R_0$). *Emerg Infect Dis*. 2019;25(1):1-4. doi:10.3201/eid2501.171901

14 Moselle, K. A., & Chang, E. (2020). CovidSIMVL – Agent-Based Modeling of Localized Transmission within a Heterogeneous Array of Locations – Motivation, Configuration and Calibration. medRxiv, (), 2020.11.01.20217943. Accessed January 06, 2021. https://doi.org/10.1101/2020.11.01.20217943.

15 He, X., Lau, E.H.Y., Wu, P. et al. (2020) Temporal dynamics in viral shedding and transmissibility of COVID-19. Nat Med 26, 672–675 (2020). https://doi.org/10.1038/s41591-020-0869-5; He, X., Lau, E.H.Y., Wu, P. *et al.* Author Correction: Temporal dynamics in viral shedding and transmissibility of COVID-19. *Nat Med* **26,** 1491–1493 (2020). https://doi.org/10.1038/s41591-020-1016-z

16 Chang, E., Moselle, K. A., & Richardson, A. (2020). CovidSIMVL --Transmission Trees, Superspreaders and Contact Tracing in Agent Based Models of Covid-19. medRxiv, (), 2020.12.21.20248673. Accessed January 06, 2021. https://doi.org/10.1101/2020.12.21.20248673.

17 Chang E. & Moselle (2021b) The anatomy of simulated Covid-19 epidemics in multiple interacting populations and spaces. Unpublished manuscript, February 5, 2021. Available on request from.
